# Brain morphometry, stimulation charge, and seizure duration in electroconvulsive therapy

**DOI:** 10.1101/2025.01.06.24319453

**Authors:** Amber M. Leaver, Chris C. Abbott, Randall T. Espinoza, Katherine L. Narr

**Affiliations:** Department of Radiology, Northwestern University, Chicago, IL, 60611; Department of Psychiatry and Biobehavioral Sciences, University of California Los Angeles, Los Angeles, CA, 90095; Department of Neurology, University of California Los Angeles, Los Angeles, CA, 90095; Department of Psychiatry, University of New Mexico; Department of Psychiatry and Behavioral Sciences, University of Washington, Seattle, WA, 98102

## Abstract

**Background:** Electroconvulsive therapy (ECT) is a well-established and effective treatment for severe depression and other conditions. Though ECT induces a generalized seizure, it is unclear why seizures are therapeutic. This study analyzed relationships between pre-treatment brain morphology, stimulation dose, and seizure duration to better understand ECT-induced seizures.

**Methods:** Pre-existing MRI data were analyzed from four cohorts with treatment refractory depression undergoing right unilateral (RUL) ECT (n=166). Regional brain morphometry and magnitude of electrical current (|E|) were analyzed, along with seizure duration and stimulation charge at seizure threshold (ECT1) and 6x seizure threshold (ECT2&3). Linear models controlled for age, sex, and cohort, corrected using false discovery rate q<0.05.

**Results:** Lower ECT1 stimulation charge correlated with less cortical surface area perpendicular to current flow, and greater |E| in nearby white matter. Lower ECT2&3 stimulation charge correlated with less cortical surface area and curvature near the temporal electrode, higher |E| in right amygdala and anterior hippocampus, and lower right thalamic and mid-hippocampal volume. Cortical surface area extending between electrodes (e.g., postcentral gyrus) positively correlated with ECT2&3 seizure duration. Successful antidepressant response associated with less cortical surface area near the temporal electrode and more |E| in anteromedial temporal lobe regions, the latter of which mediated the effects of the former on antidepressant response.

**Conclusions:** Pre-treatment brain morphology influences ECT-induced seizure, particularly in regions near ECT electrodes and those relevant to seizure initiation and modulation. Personalized dosing based on head morphology and other factors may improve antidepressant outcomes and reduce side effects.

## INTRODUCTION

Electroconvulsive therapy (ECT) is an effective treatment for severe or treatment-refractory depression and other conditions, in which carefully titrated electrical stimulation elicits brief, generalized tonic-clonic seizures to change the brain to improve symptoms (1). Neuroimaging studies of ECT have shown robust and replicable brain plasticity after treatment, including increased hippocampal volume (2–13). Some of these changes occur most in people who respond well to ECT, typically in corticolimbic and prefrontal regions already implicated in depressive neurobiology (4,11,14–16). However, it has remained unclear how seizures cause neuroplasticity leading to symptom improvement in ECT (17).

During ECT, seizure activity is monitored using 2-channel EEG to inform device settings and clinical decisions over a series of treatments (18,19). Seizure duration on EEG is routinely noted in the medical record, and electrical stimulus dose can be increased to improve seizure expression associated with better outcomes (18–20). Though some studies have linked very brief seizures with greater risk for poor antidepressant outcome in ECT (21,22), seizure duration by itself does not appear to have a strong relationship with antidepressant response (18,23). For example, studies testing low-amplitude or low-charge stimulation have elicited seizures of typical length, but were not therapeutic (19,24,25). Nevertheless, seizure duration is a widely available and used potential indicator of individual variability in seizure expression that could yield insight into how and why seizures behave differently in different people during ECT, even if its clinical utility may be unclear (18).

Variability in the electrical stimulus used to initiate seizure activity is another potential source of insight. In modern ECT, right unilateral (RUL) electrode position is typically used in a standard brief or ultra-brief pulse stimulation protocol, with one electrode near right temple, and one electrode to the right of the skull vertex (Fig 1A) (26). Pulse width and current amplitude are initially fixed, and the frequency and/or train duration of electrical pulses are varied, especially during the first session to determine individual seizure threshold, and then across subsequent sessions to maintain desired seizure expression (18). Though each constituent part of the electrical stimulus is likely to have its own independent effects on neuronal and/or seizure activity when varied across or within patients in ECT (27), an established way to summarize stimulation dose is calculating total charge in millicolumbs (mC). Stimulation charge varies across RUL and other ECT protocols; for example, ultra-brief pulse width stimulation uses less charge on average, and may associate with fewer cognitive side effects (28). Indeed, higher stimulation, whether delivered with higher amplitude or higher total charge, tends to associate with greater risk for cognitive side effects (25,28–30). Yet, within a given ECT protocol, individual variability in stimulation charge is likely to reflect how responsive each patient is to electrical seizure induction. For example, stimulation charge is associated with age and sex, with older and male patients typically requiring more stimulation (31), presumably due to differences in cortical excitability and anatomical differences (e.g., head size, skull thickness), respectively. Knowledge of how brain morphology contributes to this variability in stimulus charge could help us understand how and where seizures are initiated and/or modulated during ECT.

**Figure 1.**
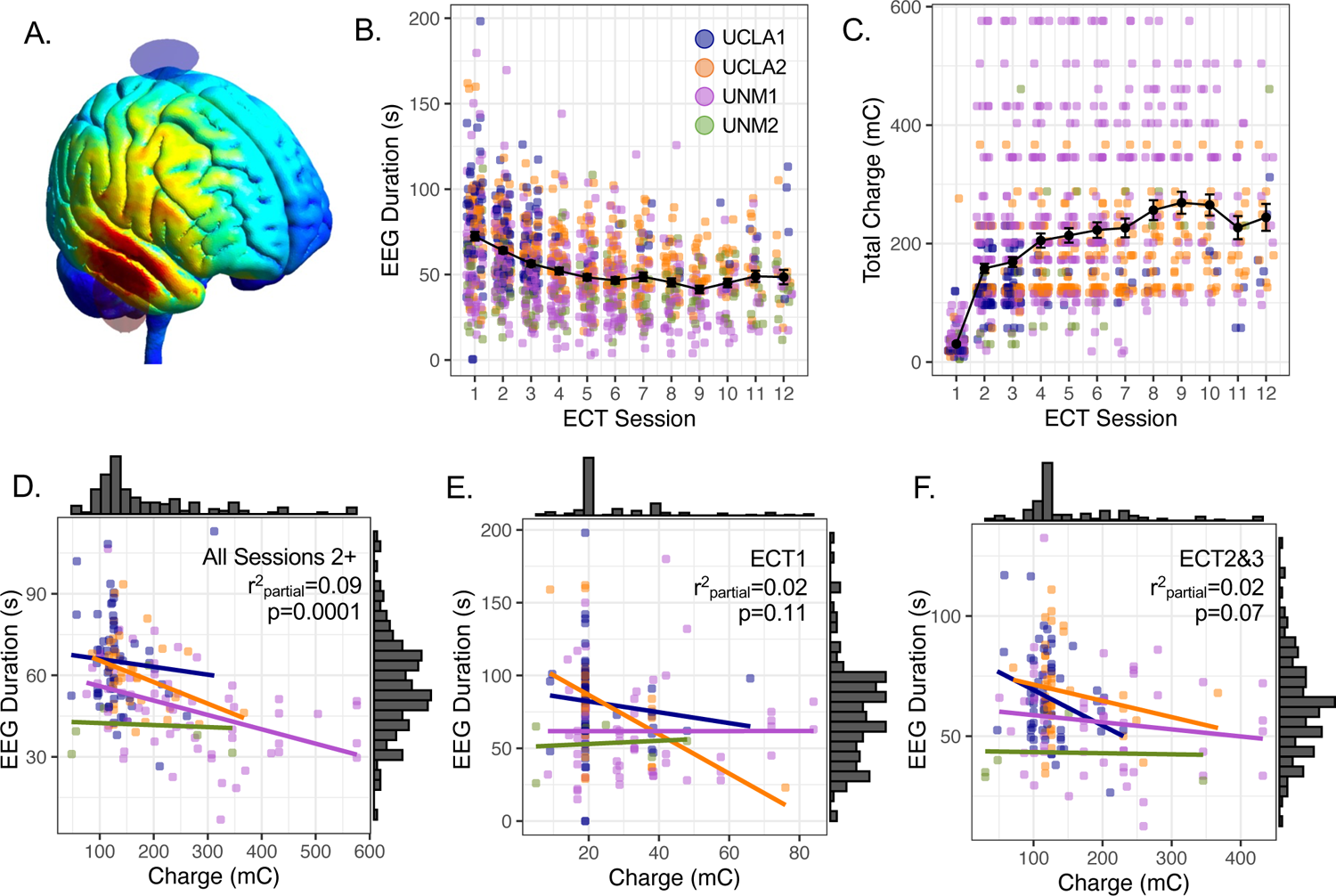
Stimulation dose and seizure duration are not strongly related during early treatment sessions. **A.** Estimated electric field magnitude, |E|, is displayed for right unilateral (RUL) ECT on an MNI template head. **B-C.** Mean seizure duration measured with EEG (sec) and mean stimulation dose in total charge (mC) are plotted over supra-threshold treatment sessions #2-#12. Error bars are standard error of the mean. **D-F.** Scatterplots relationships between seizure duration and stimulation dose during all supra-threshold sessions (D) and early treatment sessions (E-F). Color reflects study cohort, and regression lines are plotted separately for each cohort.

Relatedly, individual differences in head tissue composition and morphology determine how much electrical stimulation reaches different parts of the brain (32–35), and are also likely to impact ECT-induced seizure. In RUL ECT, electrical current magnitude, or |E|, is estimated to be highest in anterior temporal cortex and inferior frontal cortex, as well as somatomotor regions under the vertex electrode (36–38). Previous studies have reported that |E| reaching hippocampus and other brain regions correlated with the magnitude of volume increases in those same regions after ECT, though not all regions showed this pattern (38–40). Understanding how individual variability in regional |E| relates to seizure duration and stimulation charge may also offer insight into the nature of seizure activity in ECT.

This study explored relationships between pre-treatment brain morphometry, seizure duration, and electrical stimulation in patients with depression undergoing RUL ECT. We leveraged existing datasets (2,25,41,42), analyzing pre-treatment MRI data and treatment parameters collected during early ECT sessions. We hypothesized that individual variability in stimulation charge and seizure duration would correlate with morphometry and estimated |E| in regions relevant to seizure physiology, e.g., thalamus and medial temporal lobes. We targeted stimulation charge and seizure duration from the first session, where each patient’s seizure threshold is estimated using the seizure titration method (26), and during sessions two and three, where supra-threshold stimulation is delivered to elicit a therapeutic seizure. Seizure duration and stimulation charge were averaged over sessions two and three to mitigate changes in these measures across sessions for some patients (20). Taken together, these analyses sought to use MRI to improve our understanding how seizures change the brain in ECT.

## MATERIALS AND METHODS

### Participants

Existing MRI data were analyzed from four separate cohorts from University of California Los Angeles (UCLA1, UCLA2) and University of New Mexico (UNM1, UNM2). All patients (n=166) were treatment-refractory and currently experiencing a major depressive episode, and gave informed written consent to participate in these studies. Additional inclusion criteria were diagnosis of major depressive disorder (UNM & UCLA) or bipolar disorder (UCLA only). Exclusion criteria included MRI contraindications (e.g., incompatible implants, claustrophobia), other comorbid neuropsychiatric or neurological conditions, and concurrent serious illness (details can be found in source publications: (2,25,41,42)). Data selection procedures are detailed in the **Supplemental Methods**.

### ECT Sessions

ECT was administered according to the clinical procedures at each site and was not altered for research for UCLA1, UCLA2, and UNM1. For UNM2, participants were randomly assigned to different stimulation amplitudes after seizure titration (25). Apart from these differences, ECT followed similar clinical procedures, including RUL electrode positioning, ultra-brief pulse stimulation at 6x seizure threshold at the 2^nd^ session onward, 3 sessions per week, and use of short-acting anesthesia and a muscle relaxant during sessions. A subset of UNM2 participants assigned to 600mA received 1.0s pulse width stimulation. Participants who transitioned to bitemporal electrode placement were retained for the current analyses if this transition occurred after the 3^rd^ treatment.

This study targeted stimulation charge and seizure duration, taken from the first ECT session (ECT1) and averaged across the 2^nd^ and 3^rd^ sessions (ECT2&3). At ECT1, stimulation parameters or “dose” reflected seizure threshold for each participant according to the seizure titration schedules at each site. At ECT2&3, stimulation dose was supra-threshold and potentially therapeutic, while also occurring soon after baseline MRI. **Supplemental Methods** describe additional details.

### Image Acquisition & Preprocessing

T1- and T2-weighted anatomical MRI were collected before treatment at or approximately 1mm^3^ isovoxel resolution (2,10,39,42). Morphological metrics were calculated using Freesurfer 7.2.0, including volume, surface area, and curvature (43–46). Electric field magnitude (|E|, V/m) was calculated using SimNIBS 4.0 (32,47) for the true amplitude applied during treatment |E|_verum_ and at a standard amplitude in order to capture the ability of the electrical stimulus to penetrate each participant’s head (800mA, |E|_standard_). Each morphological and |E| metric was calculated and/or averaged within each brain region using standard atlases in Freesurfer (aparc.2009s, hippocampus, amygdala, thalamus) and harmonized using neuroCombat (48). Additional details in **Supplemental Methods**.

### Statistical Analyses

Statistical analyses were completed in R (49). Before MRI analyses, relationships amongst ECT parameters (i.e., seizure duration, stimulation charge, and others) were analyzed. Linear regression tested relationships between seizure duration (dependent variable) and stimulation charge (independent variable) controlling for age, sex, and cohort (regressors of no interest), and vice versa (i.e., stimulation charge as dependent variable and seizure duration as independent variable). In this way, we were able to (1) assess relationships between stimulation charge and seizure duration, and (2) explore relationships between each of these variables and age and sex. We also reported pairwise correlations amongst stimulation charge and its constituent parameters (i.e., amplitude, pulse width, frequency, paired pulse duration) separately for each cohort, because amplitude was manipulated in UNM2.

Main MRI analyses used linear regression to assess whether pre-treatment MRI statistically predicted seizure duration or stimulation charge during the first treatment (i.e., with at-threshold stimulation at ECT1) or during early therapeutic treatments (i.e., with supra-threshold stimulation at ECT2&3). In the first analysis, MRI metric was the dependent variable, and independent variables were ECT1 stimulation charge as the regressor of interest, and ECT1 seizure duration, age, sex, and cohort as additional regressors of no interest. In the second model, MRI metric was the dependent variable, ECT1 seizure duration was the regressor of interest, and ECT1 stimulation charge, age, sex, and cohort were regressors of no interest. These same models were applied again using stimulation charge and seizure duration for ECT2&3 to yield four total analyses. Because seizure duration and stimulation charge may be correlated (especially in later treatments), models assessing seizure duration included stimulation charge as a nuisance regressor of no interest, and vice versa. These models were applied to each structural MRI metric calculated by Freesurfer and (separately) to |E| calculated by SimNIBS in each atlas region, corrected at p_FDR_<0.05 within each MRI metric type. **Supplemental Methods** describe significance testing, effect size calculation, and follow-up analyses (including antidepressant response).

## RESULTS

### Treatment parameters

Stimulation charge increased on average over sessions, tracking with decreasing seizure duration as expected (**Figure 1A-B, Figure S1**). Stimulation charge and seizure duration were correlated when averaging across all supra-threshold sessions (**Figure 1C**). However, stimulation charge and seizure duration were not strongly correlated within early treatment sessions (**Table S1**), supporting their use as independent measures in MRI analyses. Seizure duration also tended to be negatively correlated with age, while charge was higher in male participants (**Table S1**). Follow-up analyses also showed that stimulation charge was not explained well by amplitude, but instead reflected a combination of multiple stimulus parameters in each cohort (**Figure S2, Supplemental Results**). Antidepressant response was also not predicted by stimulation charge or seizure duration at ECT1 or ECT2&3 (**Table S2, Figure S3, Supplemental Results**).

### Brain macro-volumes

Total cortical gray matter volume before treatment positively correlated with seizure duration and stimulation charge at ECT1 and ECT2&3 (p_uncorr_<0.05; **Table S3**). Estimated total intracranial volume before treatment also correlated with seizure duration and stimulation charge at ECT1. Subcortical gray matter volume predicted ECT2&3 stimulation charge. Effect size was largest for correlation between right cortical gray matter and seizure duration at ECT2&3, partial r^2^=0.062, but effect sizes were small overall for brain macro-volumes.

### Stimulation charge and regional brain morphometry

ECT1 charge was statistically predicted by pre-treatment surface area on the superior surface of the right lateral fissure, including frontal operculum, circular sulcus (superior insula), and posterior lateral fissure (p_FDR_<0.05; **Figure 3**, **Table 2**). Surface area of right inferior temporal cortex and on the left superior cortical surface (superior frontal gyrus, paracentral gyrus and sulcus) also correlated with ECT1 charge. Effect size was largest for right frontal operculum (partial r^2^=0.15), with full model fit (i.e., also including age, sex, and cohort) for this region explaining a larger amount of variance in ECT1 charge (adjusted r^2^=0.26).

**Figure 2.**
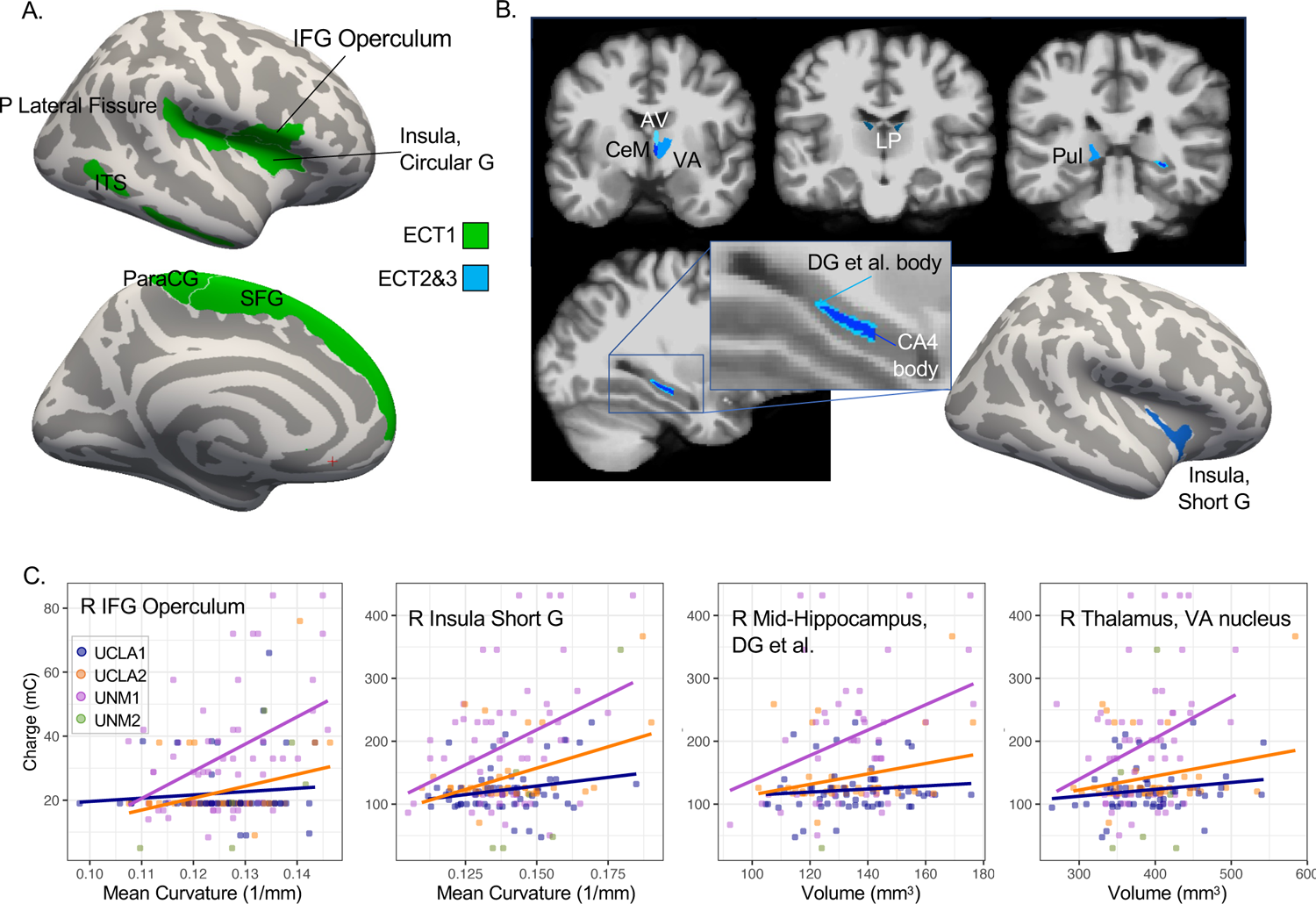
Pre-treatment regional morphometry and stimulation charge. **A.** At ECT1, stimulation charge at or near seizure threshold associated with pre-treatment morphometry in cortical regions near and between electrodes (green) displayed on template cortical surface. **B.** At ECT2&3, supra-threshold stimulation charge associated with pre-treatment morphometry in thalamic nuclei, right hippocampus body, and insula (blue). **C.** Scatter plots display positive relationships between representative morphometric features and stimulation charge in mC, with separate regression lines plotted for each cohort (except UNM2, where n=6).

**Figure 3.**
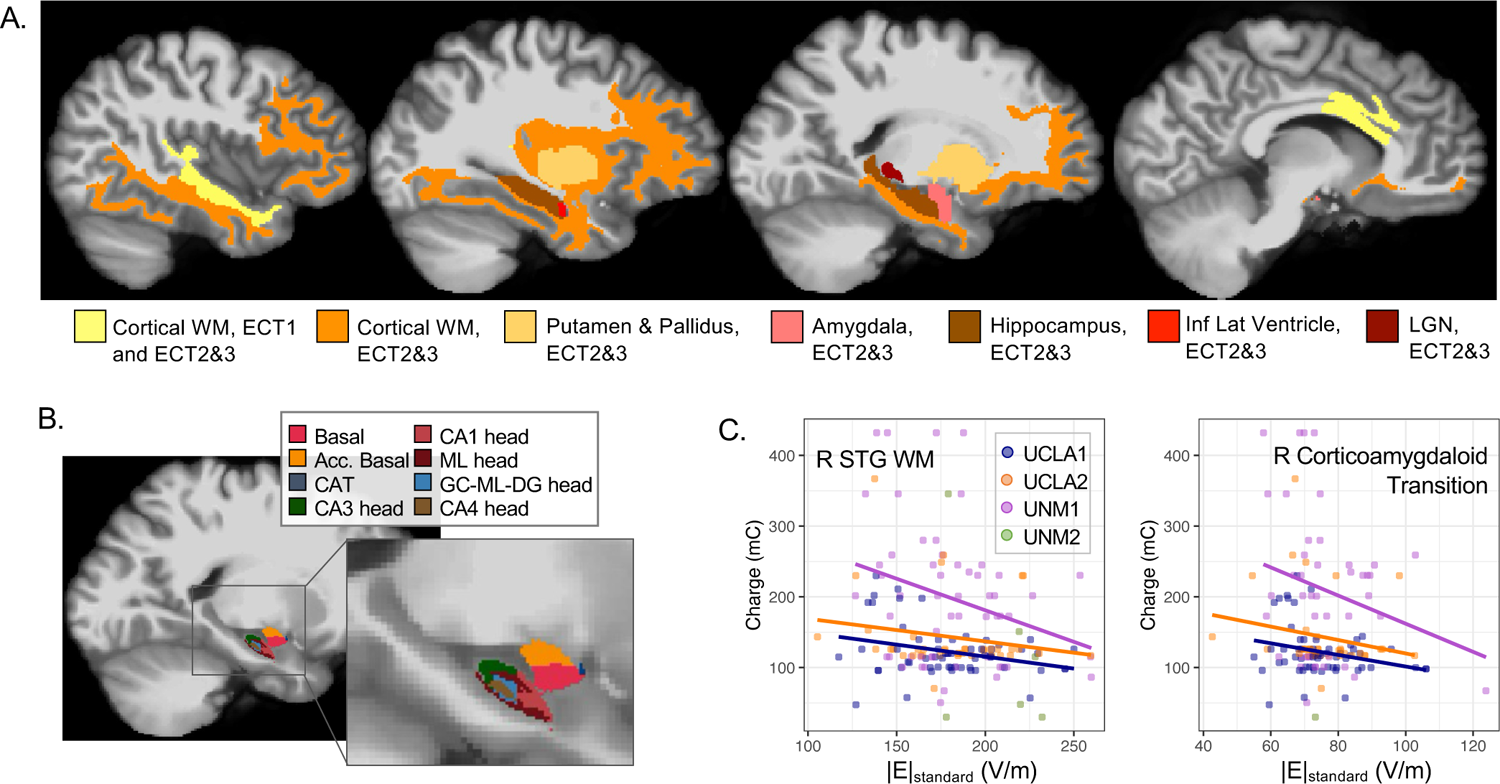
Estimated |E| and stimulation charge. **A.** Regions showing relationships between stimulation charge and both |E|_verum_ p_FDR_ < 0.05 and |E|_standard_ p < 0.05 are displayed, for ECT1 and ECT2&3 in yellow and for ECT2&3 only in warm colors. Note that |E|_standard_ reflects the ability of electrical current to penetrate the head, regardless of stimulation amplitude applied, while |E|_verum_ was calculated with stimulation amplitude used during treatment. **B.** Hippocampal subregions and amygdala nuclei showing relationships between supra-threshold stimulation charge (ECT2&3) and |E| are displayed; colors reflect the standard Freesurfer LUT for these atlases. **C.** Scatter plots display positive relationships between |E|_standard_ in V/m and stimulation charge in mC, with separate regression lines plotted for each cohort (except UNM2, where n=6).

**Table 1.** Demographic and Clinical Information by Site.

|  | UCLA 1 | UCLA 2 | UNM 1 | UNM 2 |
| --- | --- | --- | --- | --- |
| Total Sample Size | 61 | 41 | 6 | 58 |
| Age in years, mean(SD) | 43.1(14.1) | 40.1(15.9) | 63.8(10.9) | 65.3(8.4) |
| Sex, F(M) | 30(31) | 26(14) | 4(2) | 44(14) |
| Diagnosis, MDD(Bipolar) | 50(9) | 31(10) | 6(0) | 58(0) |
| ECT1 Seizure Duration in s, mean(SD) | 81.3(32.2) | 83.2(31.3) | 53.3(15.8) | 61.8(33.8) |
| ECT1 Stimulation Charge in mC, mean(SD) | 22.3(9.7) | 22.6(10.9) | 23.4(17.4) | 34.1(18.8) |
| Baseline HDRS, mean(SD) | 23.2(5.8) | 21.5(6.9) | 24.3(4.9) | 27.7(5.4) |
| Post-treatment HDRS, mean(SD) | 12.4(7.0) | 15.2(7.4) | 3.2(2.7) | 13.5(8.4) |

**Table 2.**
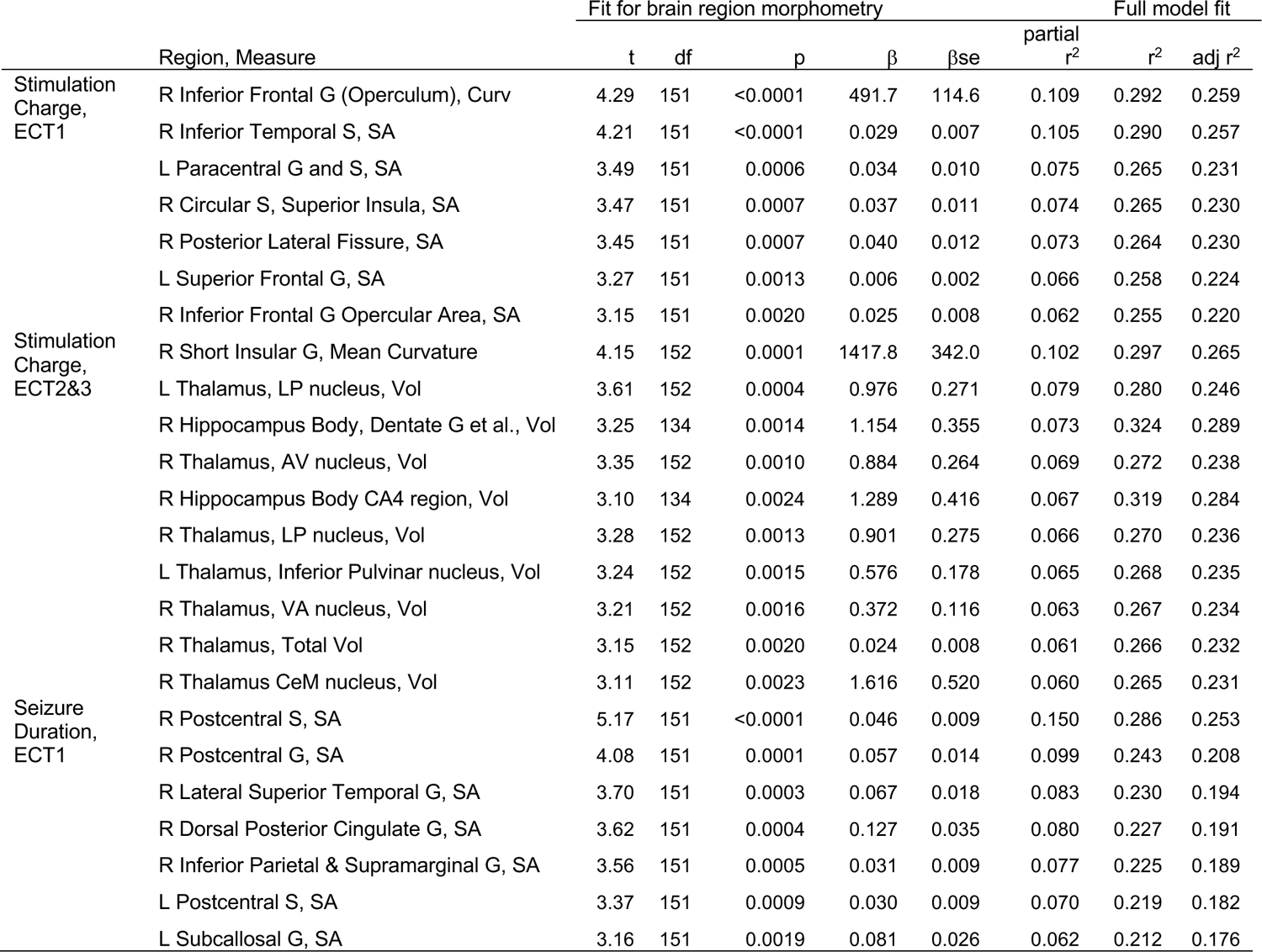
Regional morphometry, all pFDR < 0.05.

ECT2&3 stimulation charge was statistically predicted by right thalamus total volume and volume of anteromedial thalamic nuclei [right anteroventral (AV), right ventral anterior (VA), bilateral lateral posterior (LP), left inferior pulvinar]. ECT2&3 charge also correlated with volume of the body of the right dentate gyrus and CA4 region of the hippocampus, and curvature of short gyrus of the right insula, all p_FDR_<0.05 (**Figure 4**, **Table 2**). The largest amount of variance in charge was explained by the curvature of short insular gyrus (partial r^2^=0.10; full model adjusted r^2^ =0.27).

**Figure 4.**
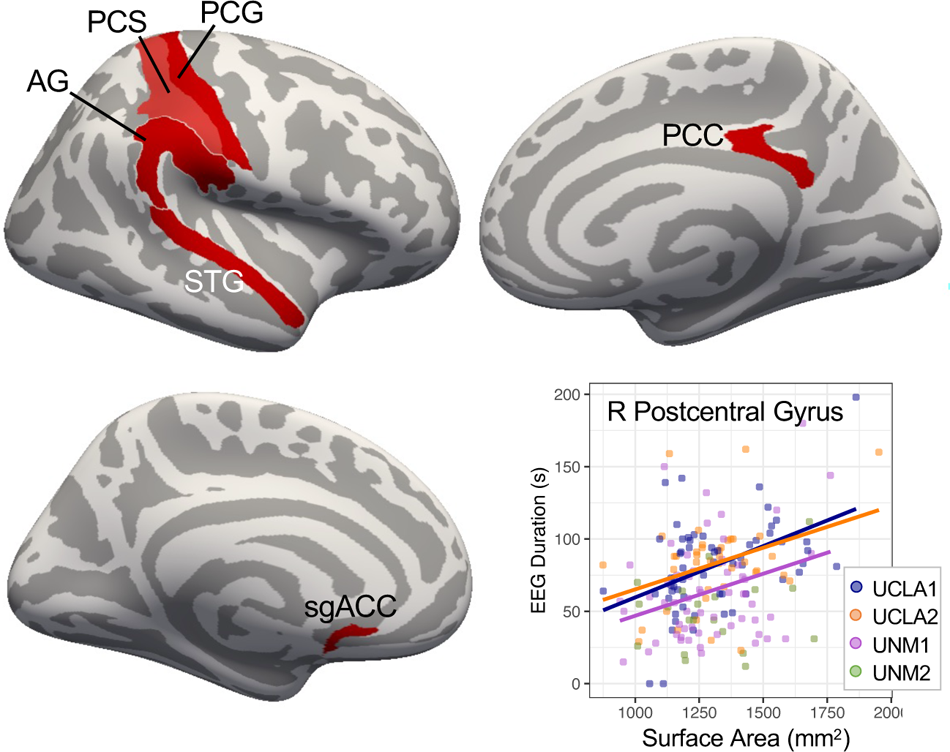
Pre-treatment cortical surface area between RUL electrodes correlates with seizure duration at first ECT session. Regions showing significant correlation (p_FDR_<0.05) between morphometry and seizure duration (ECT1) are displayed on template cortical surfaces (red). Scatter plot at bottom right displays positive relationship between a representative morphometric feature and seizure duration, with separate regression lines plotted for each cohort (except UNM2, where n=6).

In follow-up analyses, the direction of each effect persisted for number of pulses, but not amplitude, and was more likely to reach p<0.05 for pulse number (**Table S4**). In follow-up analyses for separate cohorts (UNM2, UCLA), the direction of each effect was consistent across cohorts, but was more likely to reach p<0.05 in the UNM2 cohort, where stimulation charge range was greater (**Table S5**).

### Stimulation charge and regional E-field magnitude

ECT1 charge was statistically predicted by |E| reaching white matter in right superior and inferior temporal cortex, as well as dorsal anterior cingulate cortex and corpus callosum (|E|_verum_ p_FDR_<0.05, |E|_standard_ p_uncorr_<0.05; **Figure 5**, **Table 3**). Effect sizes were modest, with |E|_standard_ in superior temporal white matter explaining the greatest amount of variance (partial r^2^=0.04; full model adjusted r^2^ =0.21).

**Figure 5.**
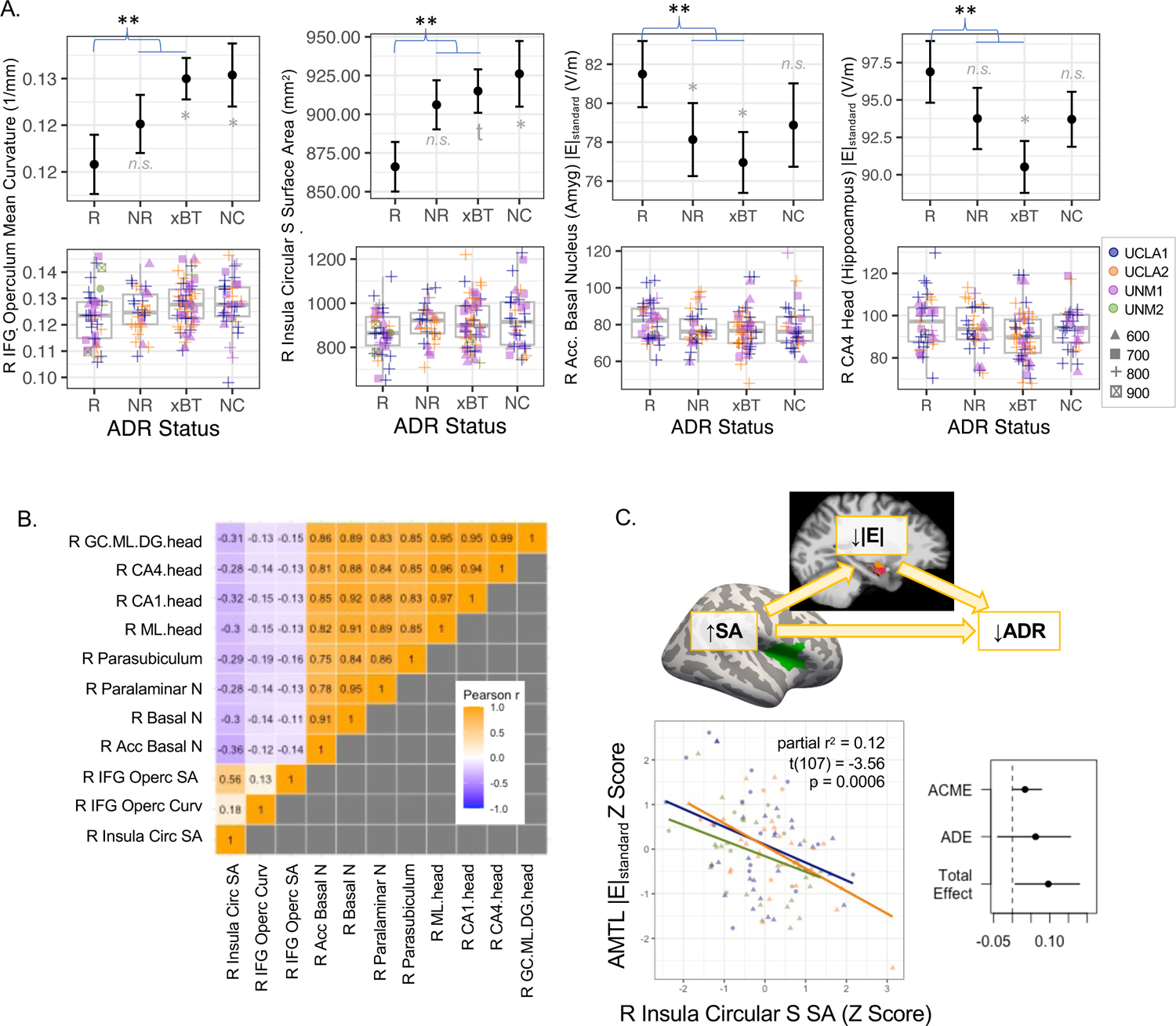
Pre-treatment morphometry and |E|_standard_ differed between people who responded to RUL ECT and nonresponders (including transition to BT ECT). **A.** Plots are displayed for representative regions showing effects of response. In top row, mean and standard error are plotted for responders (R, >50% HDRS improvement), nonresponders (NR, <50% HDRS improvement), transition to bitemporal ECT (xBT), and non-completers (NC). In second row, boxplots are displayed, along with datapoints for each participant, with color indicating cohort and shape amplitude. **B.** Cross-correlation and heat map are displayed for all regional metrics showing effects of response p < 0.05. **C.** Scatter plot displays opposing relationship between |E|_standard_ in basal nucleus of the amygdala and right anterior insula surface area. Schematic of mediation analysis is displayed at top, and mean effects are plotted for average causal mediated effect (ACME), average direct effect (ADE), and total effect; error bars are 95% confidence intervals.

**Table 3.**
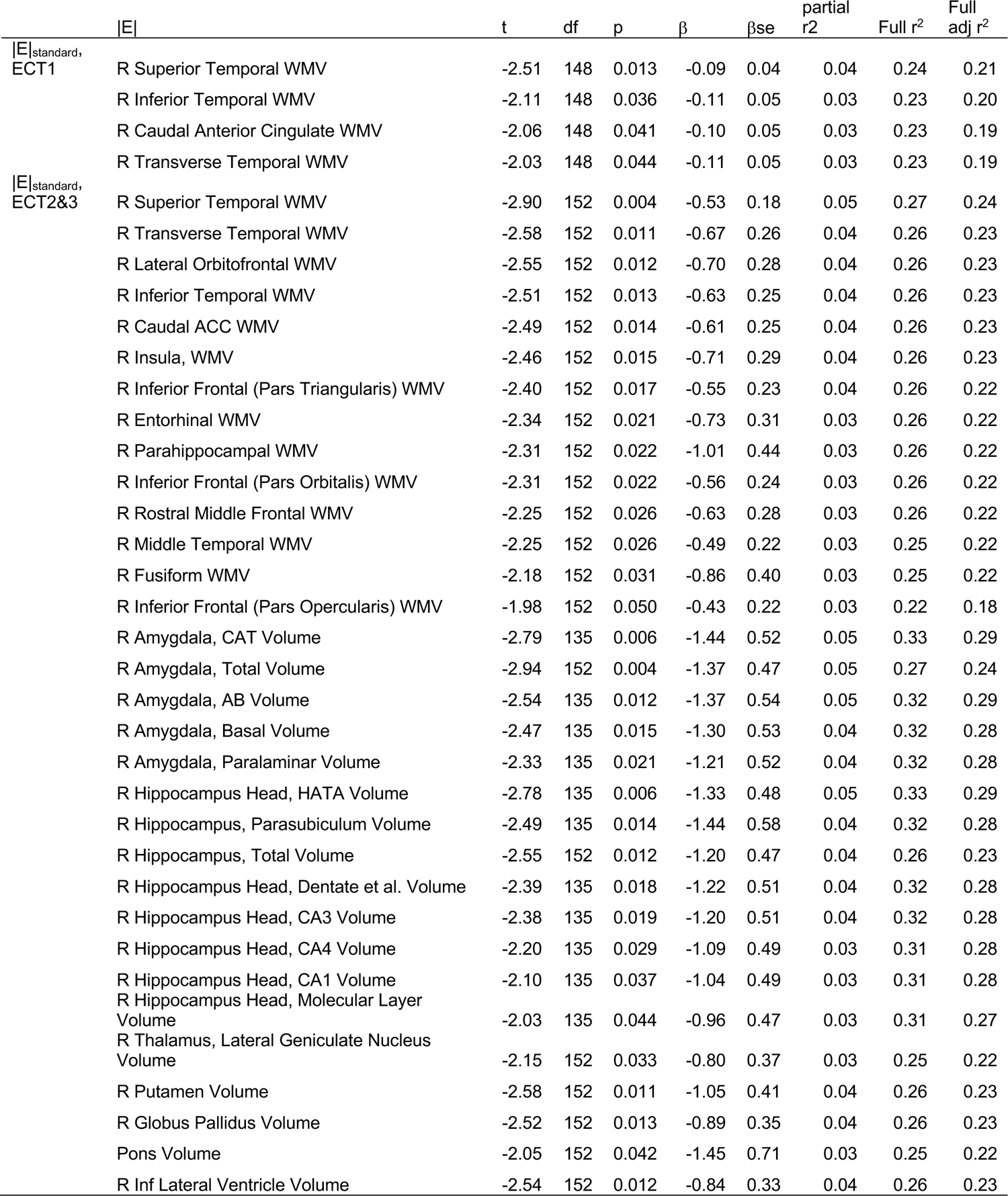
Regional estimated |E|_standard_ and Stimulation Charge (of |E|_verum_ effects p_FDR_<0.05)

Stimulation charge during ECT2&3 was statistically predicted by estimated |E| reaching several right amygdala nuclei and anterior hippocampal subregions (**Figure 5**). Stimulation charge during ECT2&3 was also associated with |E| in right putamen, globus pallidus, and white matter in right temporal, prefrontal, and insular cortex measured before treatment (|E|_verum_ p_FDR_<0.05, |E|_standard_ p_uncorr_<0.05; **Table 3**). Effect sizes were modest, with |E|_standard_ reaching the right corticoamygdaloid transition explaining the greatest amount of variance in stimulation charge (partial r^2^=0.05; full model adjusted r^2^ =0.29).

In follow-up analyses, the direction of effects from the main analysis of |E|_standard_ persisted for number of pulses, but not amplitude, and were more likely to reach p<0.05 for number of pulses (**Table S6**). The direction of each effect also remained consistent across cohorts, but was more likely to reach p<0.05 in the UCLA cohort, where a single amplitude (800 mA) was delivered across more participants (**Table S7**).

### Seizure duration, regional brain morphometry, and E-field magnitude

ECT1 seizure duration was statistically predicted by surface area of lateral cortical regions between RUL electrodes before treatment, including right postcentral gyrus and sulcus, and right lateral superior temporal, inferior parietal, and supramarginal gyri, p_FDR_<0.05 (**Figure 2**, **Table 2**). Surface area of left subcallosal cingulate gyrus, right dorsal posterior cingulate gyrus, and left postcentral sulcus also positively correlated with ECT1 seizure duration. Effect size was largest for right postcentral sulcus area (partial r^2^=0.15, full model adjusted r^2^=0.25). When analyzing ECT2&3 seizure duration, no relationships with regional brain morphometry were found. No relationships were found between seizure duration and |E|.

### Relationships with antidepressant response to RUL ECT

Follow-up analyses measured whether regions identified in the main analyses differed in people who responded to RUL ECT compared with nonresponders; nonresponders included transition to BT ECT (**Figure 6**; **Table 4**). Here, responders had less surface area on average in right anterior insula (circular sulcus) and frontal operculum, and greater curvature in right frontal operculum. |E|_standard_ was also higher for responders in anterior hippocampus and select amygdala nuclei.

**Table 4.** Main analysis effects showing differences in responders to RUL ECT, p < 0.05.

| | | t | df | p | $\beta$ | $\beta_{se}$ | partial<br>$r^2$ |
| --- | --- | --- | --- | --- | --- | --- | --- |
| Morph x<br>Charge<br>ECT1 | R Inferior Frontal G Opercular Area, SA | 2.58 | 117 | 0.011 | 64.66 | 25.08 | 0.05 |
|  | R Circular S, Superior Insula, SA | 2.01 | 117 | 0.047 | 38.211 | 18.991 | 0.033 |
|  | R Inferior Frontal G Opercular Area, Mean Curv. | 2.80 | 117 | 0.006 | 0.005 | 0.002 | 0.063 |
| \|E\|<sub>standard</sub> x<br>Charge<br>ECT2&3 | R Amygdala, Paralaminar Volume | -3.02 | 107 | 0.003 | -6.834 | 2.261 | 0.079 |
|  | R Hippocampus, Parasubiculum Volume | -2.75 | 107 | 0.007 | -5.558 | 2.019 | 0.066 |
|  | R Hippocampus Head, Molecular Layer Volume | -2.73 | 107 | 0.007 | -6.830 | 2.499 | 0.065 |
|  | R Amygdala, Basal Volume | -2.69 | 107 | 0.008 | -5.927 | 2.201 | 0.063 |
|  | R Hippocampus Head, CA1 Volume | -2.64 | 107 | 0.009 | -6.420 | 2.427 | 0.061 |
|  | R Hippocampus Head, CA4 Volume | -2.27 | 107 | 0.025 | -5.495 | 2.424 | 0.046 |
|  | R Hippocampus Head, Dentate et al. Volume | -2.16 | 107 | 0.033 | -5.044 | 2.340 | 0.042 |
|  | R Amygdala, AB Volume | -1.97 | 107 | 0.051 | -4.095 | 2.079 | 0.035 |

Examining these regions that differed between responders and nonresponders to RUL ECT, positive pairwise correlations were noted in |E|_standard_ amongst right anteromedial temporal lobe (AMTL) regions (e.g., amygdala nuclei, anterior hippocampal subregions) and in surface area amongst cortical regions. An opposing relationship was also noted between right anterior insula surface area and right AMTL |E|_standard_ (**Figure 6**), such that participants with greater right AMTL |E|_standard_ tended to have less right anterior insula surface area. In a follow-up mediation analysis, mean AMTL |E|_standard_ partially mediated (explained) the relationship between anterior insula surface area and antidepressant response (mediated effect or ACME=0.03, 95% CI=[0.001, 0.08], p=0.04).

## DISCUSSION

This study analyzed relationships between pre-treatment brain morphology, seizure duration on EEG, and the amount of stimulation required (or chosen) during RUL ECT in participants experiencing a severe, treatment-refractory major depressive episode. When stimulation was at seizure threshold (ECT1), stimulation charge correlated with surface area of cortical tissues separating RUL electrodes (i.e., perpendicular to current flow), as well as estimated magnitude of electrical current (|E|) in nearby cortical white matter. When supra-threshold doses were delivered (ECT2&3), stimulation charge correlated with tissue volume in subcortical structures, including thalamic nuclei implicated in seizure generalization and termination (50–52) and mid-hippocampus implicated in antidepressant response to ECT (53,54). ECT2&3 charge also correlated with |E| reaching seizure-genic regions in right anteromedial temporal lobe (AMTL) like amygdala (55,56), which was partially constrained by cortical surface area near the right temporal electrode and related to antidepressant response. ECT2&3 seizure duration correlated with cortical surface area extending between RUL electrodes (i.e., parallel to electrical current flow), suggesting that stimulating more neurons may lead to modestly longer seizure (57). Taken together, these results demonstrate that pre-treatment brain morphology impacts ECT-induced seizure. Improved mechanistic understanding of ECT-induced seizures, and particularly seizure variability across patients and sessions, could help develop approaches to precision dosing and other improvements to ECT.

### Subcortical vs. Cortical Involvement in ECT-Induced Seizure

Stimulation amplitude, and therefore |E|, typically does not change between seizure titration and subsequent sessions. However, the number, width, and/or frequency of electrical pulses (and therefore stimulation charge) delivered across the |E| distribution does change. In our study, ECT1 charge correlated exclusively with cortical metrics, while ECT2&3 charge correlated with subcortical metrics. Though qualitative, this difference is notable given that each pulse delivered also likely exceeds the threshold for neuronal firing in a majority of brain tissue (58,59). This could suggest that seizure initiation occurs primarily in right cortical regions when stimulation is at seizure threshold and pulse trains are shorter (though subcortical structures like thalamus are almost certainly recruited during these generalized seizures (60)), while subcortical regions like thalamus, hippocampus, and amygdala become more relevant to ECT-induced seizure during supra-threshold stimulation, i.e., when pulse trains are longer. Correlations noted between ECT2&3 pulse number and |E|_standard_ or morphometry in subcortical regions also support this idea. Given the seizure-genic nature of amygdala and nearby anteromedial temporal lobe regions (55,61), we speculate that |E| reaching anteromedial temporal lobe (AMTL) regions may only be sufficient to initiate seizure activity in AMTL during supra-threshold sessions, where longer pulse trains are applied. However, studies directly measuring the distribution of seizure activity as it unfolds during at- and supra-threshold ECT with adequate spatial and temporal resolution are needed to confirm this interpretation.

If the site(s) of seizure initiation differ for at-versus supra-threshold dosing in RUL ECT, this could have implications for the seizure titration method as a means of determining threshold. For example, if antidepressant outcome in RUL ECT relies on the brain’s response to seizure activity initiated in right AMTL, more precisely matching seizure threshold in right AMTL in each patient could improve outcomes (i.e., versus matching seizure threshold in right cortical regions, presumably as in current seizure titration methods). Conversely, if the location(s) of seizure initiation is not relevant to antidepressant response to RUL ECT, then perhaps a more general measure of cortical excitability could offer an equally accurate but less invasive/burdensome means to estimate seizure threshold (e.g., TMS-induced MEPs, or a functional neuroimaging-based biomarker). Future studies could consider these issues to improve seizure threshold estimation, to more precisely determine dose without seizure and improve outcomes.

ECT2&3 charge also correlated with pre-treatment anteromedial thalamic and mid-hippocampal volume but not |E|, so perhaps these effects are more relevant to seizure generalization than initiation. The thalamus is well-known to play a role in seizure generalization and modulation (50,60) (62–68), and thalamic deep brain stimulation can be effective in treating refractory epilepsy (51,52). ECT-MRI studies also report correlation between increased thalamic CBF after the first supra-threshold session and later antidepressant outcome (UCLA1 (53)), as well as thalamic plasticity after ECT (69,70). Changes in mid-hippocampal function also associate with antidepressant response in ECT (UCLA1 (53,54)), and connections between hippocampus and anteromedial thalamus are thought to be involved in modulating hippocampal seizure (71,72). Future studies examining connectivity amongst these regions in relation to ECT2&3 charge or seizure expression (e.g., ictal theta (73)) could be informative in predicting susceptibility to generalized seizure in ECT.

### Cortical Morphology, Current Flow, and Antidepressant Response to ECT

In this study, ECT1 charge positively correlated with the size of cortical surfaces extending perpendicular to electrical current flow. This could suggest that patients with larger cortical surfaces obstructing the flow of electrical current between electrodes required more stimulation to elicit seizures during titration, perhaps due to greater shunting of electrical current in less-resistive CSF across cortical surfaces perpendicular to current flow (74). This interpretation is also supported by our finding that patients with less |E| penetrating nearby right cortical white matter also tended to require greater ECT1 charge (perhaps) to compensate. Taken together, these results demonstrate how the size (and/or shape) of cortex near electrodes may constrain stimulus delivery to key regions during ECT.

Anterior insula surface area near the right temporal electrode also appeared to influence how much stimulation reached right amygdala and anterior hippocampus (i.e., AMTL |E|_standard_), both of which also associated with antidepressant response to RUL ECT. Here, nonresponders (including transition to BT ECT) tended to have larger right insula surface area and less |E|_standard_ in right amygdala and anterior hippocampus. Though previous studies reported finding no relationship between antidepressant response and hippocampal or amygdalar |E| (38,39), these studies did not analyze hippocampal subregions or amygdala nuclei and/or define transition to BT ECT as nonresponse to RUL ECT. Our results suggest that adjusting dose to increase |E| in right AMTL (e.g., greater amplitude, more pulses, or adjusting electrode position) could improve outcomes for some patients in RUL ECT. However, it is also important to note that effect sizes were relatively modest, and other factors are likely to play a role in ECT-induced seizure, antidepressant response, and stimulus delivery (e.g., skull thickness).

### Morphological predictors of seizure duration

Patients with longer seizure duration at ECT2&3 tended to have more cortical surface area extending between RUL electrodes, including right postcentral sulcus and gyrus, superior temporal gyrus, and adjacent parietal regions. These same brain regions extend parallel with the direction of current flow, and also tended to receive greater |E| than elsewhere in the brain (**Fig 1A**; (35,36,57)). Cortical surface area is linked with a number of ontological/developmental processes (75,76), and could reflect the number or size of cortical columns within a given region (77,78). Thus, the present results could suggest that the number neurons maximally stimulated during pulsed electrical current may contribute to seizure length during typical RUL treatment sessions (57). Also of note, subgenual and posterior cingulate was associated with seizure duration but not antidepressant response, despite their roles in depressive neurobiology (79,81,83) (**Supplemental Discussion**).

These results also suggest that brain morphology could inform statistical models predicting seizure length; the combination of age, sex, and right precentral sulcus area explained 25% of the variance in seizure duration, a large effect size (Table 2). It is unclear whether seizures that are too brief are undesirable, as some studies report cases where very brief seizures were therapeutic (18). However, predictive models that identify risk for long seizures associated with side effects (80) could be useful. Yet, seizures were of typical length in this study, and different aspects of brain morphology and/or function may better explain seizure length (e.g., measures of neuronal excitability, connectivity, excitatory/inhibitory neurotransmitter balance, or other factors). A recent study noted a modest correlation between increased MTL volume and summed durations of all seizures during ECT course; however, it is difficult to dissociate from the number of treatments in each patient’s treatment course from this summed metric (40). Regardless of the clinical utility of predictive models for seizure length, future studies seeking to further explain variance in seizure duration could still yield insight into the neurobiological nature of therapeutic seizure in ECT.

## Conclusions

This study identified instances where brain morphology influenced ECT-induced seizure. Seizures were assessed indirectly using the amount of electrical stimulation given (charge), the amount of stimulation reaching different brain areas (|E|), and seizure duration. Though we used these data to infer how seizure activity may progress through the brain during ECT, we have not measured this directly, an important limitation to consider. However, ictal neuroimaging also has limitations to consider: molecular imaging lacks temporal resolution, scalp EEG lacks spatial resolution (particularly in deep MTL structures), and intracranial EEG lacks whole-brain coverage. As such, there is immense value in retrospective analysis of existing ECT-MRI datasets (82), and converging evidence across approaches will ultimately be what propels the field forward. Additional limitations of the current study include the reliability (84) and/or fidelity of morphometric atlases used (e.g., cerebellum, piriform, brainstem nuclei, and other structures were omitted here), cross-site differences in clinical procedures, and other factors. Nevertheless, we expect these findings will inform future efforts to test (or refute) these ideas (e.g., whether site(s) of seizure initiation differ during seizure titration vs. supra-threshold stimulation, whether brain morphology can be used in models of personalized dosing).

## Data Availability

The majority of this data are available at the NIMH Data Archive. All reasonable requests for other data not available in the NDA will be considered on a case-by-case basis by the individual authors responsible for each dataset (due to the sensitivity of the data).

## ACKNOWLEDGEMENTS

This work was supported by the NIH, including R03 MH121769 to Dr. Leaver, R01 MH092301 and U01 MH110008 to Drs. Narr and Espinoza, P20 GM103472 and U01 MH111826 to Dr. Abbott. This content is solely the responsibility of the authors and does not necessarily reflect the official views of the National Institutes of Health. This work was also supported in part through the computational resources and staff contributions provided for the Quest high performance computing facility at Northwestern University which is jointly supported by the Office of the Provost, the Office for Research, and Northwestern University Information Technology. This work was also supported in part by the Muriel Harris Chair of Geriatric Psychiatry at UCLA to Dr. Espinoza.

## CONFLICT OF INTEREST

All authors declare no conflicts of interest.

## Notes

### Competing Interest Statement

The authors have declared no competing interest.

### Author Declarations

Northwestern University Institutional Review Board has approved a waiver for this research, stating that the research is exempt from review under category 4 (secondary research on data or specimens).

## REFERENCES

1. Espinoza RT, Kellner CH (2022): Electroconvulsive Therapy. N Engl J Med 386: 667–672.

2. Joshi SH, Espinoza RT, Pirnia T, Shi J, Wang Y, Ayers B, et al. (2015): Structural plasticity of the hippocampus and amygdala induced by electroconvulsive therapy in major depression. Biol Psychiatry 79: 282–92.

3. Tendolkar I, Beek M, Oostrom I, Mulder M, Janzing J, Voshaar RO, Eijndhoven P (2013): Electroconvulsive therapy increases hippocampal and amygdala volume in therapy refractory depression: a longitudinal pilot study. Psychiatry Res 214: 197–203.

4. Redlich R, Opel N, Grotegerd D, Dohm K, Zaremba D, Bürger C, et al. (2016): Prediction of Individual Response to Electroconvulsive Therapy via Machine Learning on Structural Magnetic Resonance Imaging Data. JAMA Psychiatry 73: 557.

5. Dukart J, Regen F, Kherif F, Colla M, Bajbouj M, Heuser I, et al. (2014): Electroconvulsive therapy-induced brain plasticity determines therapeutic outcome in mood disorders. Proc Natl Acad Sci U S A 111: 1156–61.

6. Cano M, Martínez-Zalacaín I, Bernabéu-Sanz Á, Contreras-Rodríguez O, Hernández-Ribas R, Via E, et al. (2017): Brain volumetric and metabolic correlates of electroconvulsive therapy for treatment-resistant depression: a longitudinal neuroimaging study. Transl Psychiatry 7: e1023.

7. Chen F, Madsen TM, Wegener G, Nyengaard JR (2009): Repeated electroconvulsive seizures increase the total number of synapses in adult male rat hippocampus. Eur Neuropsychopharmacol 19: 329–338.

8. Madsen TM, Treschow A, Bengzon J, Bolwig TG, Lindvall O, Tingström A (2000): Increased neurogenesis in a model of electroconvulsive therapy. Biol Psychiatry 47: 1043–1049.

9. Perera TD, Coplan JD, Lisanby SH, Lipira CM, Arif M, Carpio C, et al. (2007): Antidepressant-induced neurogenesis in the hippocampus of adult nonhuman primates. J Neurosci Off J Soc Neurosci 27: 4894–4901.

10. Abbott CC, Jones T, Lemke NT, Gallegos P, McClintock SM, Mayer AR, et al. (2014): Hippocampal structural and functional changes associated with electroconvulsive therapy response. Transl Psychiatry 4: e483.

11. Leaver AM, Espinoza R, Pirnia T, Joshi SH, Woods RP, Narr KL (2016): Modulation of Intrinsic Brain Activity by Electroconvulsive Therapy in Major Depression. Biol Psychiatry Cogn Neurosci Neuroimaging 1: 77–86.

12. Argyelan M, Lencz T, Kaliora S, Sarpal DK, Weissman N, Kingsley PB, et al. (2016): Subgenual cingulate cortical activity predicts the efficacy of electroconvulsive therapy. Transl Psychiatry 6: e789.

13. Njau S, Joshi SH, Leaver AM, Vasavada M, Fleet J, Espinoza R, Narr KL (2016): Variations in myo-inositol in fronto-limbic regions and clinical response to electroconvulsive therapy in major depression. J Psychiatr Res 80: 45–51.

14. Mulders PCR, Llera A, Beckmann CF, Vandenbulcke M, Stek M, Sienaert P, et al. (2020): Structural changes induced by electroconvulsive therapy are associated with clinical outcome. Brain Stimulat 13: 696–704.

15. Leaver AM, Wade B, Vasavada M, Hellemann G, Joshi SH, Espinoza R, Narr KL (2018): Fronto-Temporal Connectivity Predicts ECT Outcome in Major Depression. Front Psychiatry 9. 10.3389/fpsyt.2018.00092

16. Pirnia T, Joshi SH, Leaver AM, Vasavada M, Njau S, Woods RP, et al. (2016): Electroconvulsive therapy and structural neuroplasticity in neocortical, limbic and paralimbic cortex. Transl Psychiatry 6: e832.

17. Deng Z-D, Robins PL, Regenold W, Rohde P, Dannhauer M, Lisanby SH (2024): How electroconvulsive therapy works in the treatment of depression: is it the seizure, the electricity, or both? Neuropsychopharmacology 49: 150–162.

18. Francis-Taylor R, Ophel G, Martin D, Loo C (2020): The ictal EEG in ECT: A systematic review of the relationships between ictal features, ECT technique, seizure threshold and outcomes. Brain Stimulat 13: 1644–1654.

19. Sackeim HA, Devanand DP, Prudic J (1991): Stimulus intensity, seizure threshold, and seizure duration: impact on the efficacy and safety of electroconvulsive therapy. Psychiatr Clin North Am 14: 803–843.

20. Luccarelli J, McCoy TH, Seiner SJ, Henry ME (2021): Changes in seizure duration during acute course electroconvulsive therapy. Brain Stimulat 14: 941–946.

21. Nobler MS, Sackeim HA, Solomou M, Luber B, Devanand DP, Prudic J (1993): EEG manifestations during ECT: effects of electrode placement and stimulus intensity. Biol Psychiatry 34: 321–330.

22. Gillving C, Ekman CJ, Hammar Å, Landén M, Lundberg J, Movahed Rad P, et al. (2024): Seizure Duration and Electroconvulsive Therapy in Major Depressive Disorder. JAMA Netw Open 7: e2422738.

23. Folkerts H (1996): The ictal electroencephalogram as a marker for the efficacy of electroconvulsive therapy. Eur Arch Psychiatry Clin Neurosci 246: 155–164.

24. Sackeim HA, Prudic J, Devanand DP, Kiersky JE, Fitzsimons L, Moody BJ, et al. (1993): Effects of stimulus intensity and electrode placement on the efficacy and cognitive effects of electroconvulsive therapy. N Engl J Med 328: 839–846.

25. Abbott CC, Quinn D, Miller J, Ye E, Iqbal S, Lloyd M, et al. (2021): Electroconvulsive Therapy Pulse Amplitude and Clinical Outcomes. Am J Geriatr Psychiatry 29: 166–178.

26. Kellner CH, Pritchett JT, Beale MD, Coffey CE (1997): Handbook of ECT. Washington, D.C.: American Psychiatric Press.

27. Peterchev AV, Rosa MA, Deng Z-D, Prudic J, Lisanby SH (2010): Electroconvulsive therapy stimulus parameters: rethinking dosage. J ECT 26: 159–174.

28. McClintock SM, Choi J, Deng Z-D, Appelbaum LG, Krystal AD, Lisanby SH (2014): Multifactorial Determinants of the Neurocognitive Effects of Electroconvulsive Therapy. J ECT 30: 165–176.

29. Sackeim HA, Prudic J, Fuller R, Keilp J, Lavori PW, Olfson M (2007): The Cognitive Effects of Electroconvulsive Therapy in Community Settings [no. 1]. Neuropsychopharmacology 32: 244–254.

30. Youssef NA, Sidhom E (2017): Feasibility, safety, and preliminary efficacy of Low Amplitude Seizure Therapy (LAP-ST): A proof of concept clinical trial in man. J Affect Disord 222: 1–6.

31. Luccarelli J, McCoy TH, Seiner SJ, Henry ME (2020): Charge required to induce a seizure during initial dose titration using right unilateral brief pulse electroconvulsive therapy. Brain Stimul Basic Transl Clin Res Neuromodulation 13: 1504–1506.

32. Saturnino GB, Puonti O, Nielsen JD, Antonenko D, Madsen KH, Thielscher A (2019): SimNIBS 2.1: A Comprehensive Pipeline for Individualized Electric Field Modelling for Transcranial Brain Stimulation. In: Makarov S, Horner M, Noetscher G, editors. Brain and Human Body Modeling: Computational Human Modeling at EMBC 2018. Cham (CH): Springer. Retrieved January 22, 2021, from http://www.ncbi.nlm.nih.gov/books/NBK549569/

33. Nielsen JD, Madsen KH, Puonti O, Siebner HR, Bauer C, Madsen CG, et al. (2018): Automatic skull segmentation from MR images for realistic volume conductor models of the head: Assessment of the state-of-the-art. NeuroImage 174: 587–598.

34. Dannhauer M, Lanfer B, Wolters CH, Knösche TR (2011): Modeling of the human skull in EEG source analysis. Hum Brain Mapp 32: 1383–1399.

35. Lee WH, Deng Z-D, Kim T-S, Laine AF, Lisanby SH, Peterchev AV (2012): Regional electric field induced by electroconvulsive therapy in a realistic finite element head model: Influence of white matter anisotropic conductivity. NeuroImage 59: 2110–2123.

36. Deng Z-D, Lisanby SH, Peterchev AV (2011): Electric field strength and focality in electroconvulsive therapy and magnetic seizure therapy: a finite element simulation study. J Neural Eng 8: 016007.

37. Fridgeirsson EA, Deng Z-D, Denys D, van Waarde JA, van Wingen GA (2021): Electric field strength induced by electroconvulsive therapy is associated with clinical outcome. NeuroImage Clin 30: 102581.

38. Argyelan M, Oltedal L, Deng Z-D, Wade B, Bikson M, Joanlanne A, et al. (2019): Electric field causes volumetric changes in the human brain. eLife 8: e49115.

39. Deng Z-D, Argyelan M, Miller J, Quinn DK, Lloyd M, Jones TR, et al. (2022): Electroconvulsive therapy, electric field, neuroplasticity, and clinical outcomes. Mol Psychiatry 27: 1676–1682.

40. Takamiya A, Bouckaert F, Laroy M, Blommaert J, Radwan A, Khatoun A, et al. (2021): Biophysical mechanisms of electroconvulsive therapy-induced volume expansion in the medial temporal lobe: A longitudinal in vivo human imaging study. Brain Stimulat 14: 1038–1047.

41. Abbott CC, Jones T, Lemke NT, Gallegos P, McClintock SM, Mayer AR, et al. (2014): Hippocampal structural and functional changes associated with electroconvulsive therapy response. Transl Psychiatry 4: e483.

42. Tozzi L, Anene ET, Gotlib IH, Wintermark M, Kerr AB, Wu H, et al. (2021): Convergence, preliminary findings and future directions across the four human connectome projects investigating mood and anxiety disorders. NeuroImage 245: 118694.

43. Destrieux C, Fischl B, Dale A, Halgren E (2010): Automatic parcellation of human cortical gyri and sulci using standard anatomical nomenclature. NeuroImage 53: 1–15.

44. Fischl B, Salat DH, Busa E, Albert M, Dieterich M, Haselgrove C, et al. (2002): Whole brain segmentation: automated labeling of neuroanatomical structures in the human brain. Neuron 33: 341–355.

45. Fischl B, Dale AM (2000): Measuring the thickness of the human cerebral cortex from magnetic resonance images. Proc Natl Acad Sci 97: 11050–11055.

46. Dale AM, Fischl B, Sereno MI (1999): Cortical surface-based analysis. I. Segmentation and surface reconstruction. NeuroImage 9: 179–194.

47. Thielscher A, Antunes A, Saturnino GB (2015): Field modeling for transcranial magnetic stimulation: A useful tool to understand the physiological effects of TMS? 2015 37th Annual International Conference of the IEEE Engineering in Medicine and Biology Society (EMBC) 222–225.

48. Fortin J-P, Parker D, Tunç B, Watanabe T, Elliott MA, Ruparel K, et al. (2017): Harmonization of multi-site diffusion tensor imaging data. NeuroImage 161: 149–170.

49. R Core Team (n.d.): R: The R Project for Statistical Computing. Vienna, Austria. Retrieved from https://www.r-project.org/

50. Mirski MA, Rossell LA, Terry JB, Fisher RS (1997): Anticonvulsant effect of anterior thalamic high frequency electrical stimulation in the rat. Epilepsy Res 28: 89–100.

51. Salanova V, Witt T, Worth R, Henry TR, Gross RE, Nazzaro JM, et al. (2015): Long-term efficacy and safety of thalamic stimulation for drug-resistant partial epilepsy. Neurology 84: 1017–1025.

52. Fisher R, Salanova V, Witt T, Worth R, Henry T, Gross R, et al. (2010): Electrical stimulation of the anterior nucleus of thalamus for treatment of refractory epilepsy. Epilepsia 51: 899–908.

53. Leaver AM, Vasavada M, Joshi SH, Wade B, Woods RP, Espinoza R, Narr KL (2019): Mechanisms of Antidepressant Response to Electroconvulsive Therapy Studied With Perfusion Magnetic Resonance Imaging. Biol Psychiatry 85: 466–476.

54. Leaver AM, Vasavada M, Kubicki A, Wade B, Loureiro J, Hellemann G, et al. (2020): Hippocampal subregions and networks linked with antidepressant response to electroconvulsive therapy. Mol Psychiatry 1–12.

55. Gale K (1992): Subcortical Structures and Pathways Involved in Convulsive Seizure Generation. J Clin Neurophysiol 9: 264–277.

56. Bertram EH (2009): Temporal lobe epilepsy: Where do the seizures really begin? Epilepsy Behav 14: 32–37.

57. Bai S, Loo C, Al Abed A, Dokos S (2012): A computational model of direct brain excitation induced by electroconvulsive therapy: comparison among three conventional electrode placements. Brain Stimulat 5: 408– 421.

58. Lee WH, Lisanby SH, Laine AF, Peterchev AV (2016): Comparison of electric field strength and spatial distribution of electroconvulsive therapy and magnetic seizure therapy in a realistic human head model. Eur Psychiatry J Assoc Eur Psychiatr 36: 55–64.

59. Abbott CC, Miller J, Farrar D, Argyelan M, Lloyd M, Squillaci T, et al. (2024): Amplitude-determined seizure-threshold, electric field modeling, and electroconvulsive therapy antidepressant and cognitive outcomes. Neuropsychopharmacology 49: 640–648.

60. Lindquist BE, Timbie C, Voskobiynyk Y, Paz JT (2023): Thalamocortical circuits in generalized epilepsy: Pathophysiologic mechanisms and therapeutic targets. Neurobiol Dis 181: 106094.

61. Piredda S, Gale K (1985): A crucial epileptogenic site in the deep prepiriform cortex. Nature 317: 623–625.

62. Enev M, McNally KA, Varghese G, Zubal IG, Ostroff RB, Blumenfeld H (2007): Imaging onset and propagation of ECT-induced seizures. Epilepsia 48: 238–244.

63. Blumenfeld H, Westerveld M, Ostroff RB, Vanderhill SD, Freeman J, Necochea A, et al. (2003): Selective frontal, parietal, and temporal networks in generalized seizures. NeuroImage 19: 1556–1566.

64. Blumenfeld H, McNally KA, Ostroff RB, Zubal IG (2003): Targeted prefrontal cortical activation with bifrontal ECT. Psychiatry Res 123: 165–170.

65. Blumenfeld H, Varghese GI, Purcaro MJ, Motelow JE, Enev M, McNally KA, et al. (2009): Cortical and subcortical networks in human secondarily generalized tonic–clonic seizures. Brain 132: 999–1012.

66. Takano H, Motohashi N, Uema T, Ogawa K, Ohnishi T, Nishikawa M, Matsuda H (2011): Differences in cerebral blood flow between missed and generalized seizures with electroconvulsive therapy: A positron emission tomographic study. Epilepsy Res 97: 225–228.

67. Takano H, Motohashi N, Uema T, Ogawa K, Ohnishi T, Nishikawa M, et al. (2007): Changes in regional cerebral blood flow during acute electroconvulsive therapy in patients with depression: Positron emission tomographic study. Br J Psychiatry 190: 63–68.

68. Bajc M, Medved V, Basic M, Topuzovic N, Babic D, Ivancevic D (1989): Acute effect of electroconvulsive therapy on brain perfusion assessed by Tc99m-hexamethylpropyl-eneamineoxim and single photon emission computed tomography. Acta Psychiatr Scand 80: 421–426.

69. Takamiya A, Kishimoto T, Liang K, Terasawa Y, Nishikata S, Tarumi R, et al. (2019): Thalamic volume, resting-state activity, and their association with the efficacy of electroconvulsive therapy. J Psychiatr Res 117: 135–141.

70. Wei Q, Bai T, Brown EC, Xie W, Chen Y, Ji G, et al. (2020): Thalamocortical connectivity in electroconvulsive therapy for major depressive disorder. J Affect Disord 264: 163–171.

71. Fei F, Wang X, Xu C, Shi J, Gong Y, Cheng H, et al. (2022): Discrete subicular circuits control generalization of hippocampal seizures. Nat Commun 13: 5010.

72. Yu T, Wang X, Li Y, Zhang G, Worrell G, Chauvel P, et al. (2018): High-frequency stimulation of anterior nucleus of thalamus desynchronizes epileptic network in humans. Brain 141: 2631–2643.

73. Miller J, Jones T, Upston J, Deng Z-D, McClintock SM, Erhardt E, et al. (2023): Electric Field, Ictal Theta Power, and Clinical Outcomes in Electroconvulsive Therapy. Biol Psychiatry Cogn Neurosci Neuroimaging 8: 760–767.

74. Nathan SS, Sinha SR, Gordon B, Lesser RP, Thakor NV (1993): Determination of current density distributions generated by electrical stimulation of the human cerebral cortex. Electroencephalogr Clin Neurophysiol 86: 183– 192.

75. Chen C-H, Gutierrez ED, Thompson W, Panizzon MS, Jernigan TL, Eyler LT, et al. (2012): Hierarchical genetic organization of human cortical surface area. Science 335: 1634–1636.

76. Wierenga LM, Langen M, Oranje B, Durston S (2014): Unique developmental trajectories of cortical thickness and surface area. NeuroImage 87: 120–126.

77. Rakic P (2009): Evolution of the neocortex: a perspective from developmental biology. Nat Rev Neurosci 10: 724–735.

78. Malach R (1994): Cortical columns as devices for maximizing neuronal diversity. Trends Neurosci 17: 101–104.

79. Leaver AM, Espinoza R, Joshi SH, Vasavada M, Njau S, Woods RP, Narr KL (2016): Desynchronization and Plasticity of Striato-frontal Connectivity in Major Depressive Disorder. Cereb Cortex N Y N 1991 26: 4337–4346.

80. Cristancho MA, Alici Y, Augoustides JG, O’Reardon JP (2008): Uncommon but serious complications associated with electroconvulsive therapy: Recognition and management for the clinician. Curr Psychiatry Rep 10: 474–480.

81. Mayberg HS, Liotti M, Brannan SK, McGinnis S, Mahurin RK, Jerabek PA, et al. (1999): Reciprocal Limbic-Cortical Function and Negative Mood: Converging PET Findings in Depression and Normal Sadness. Am J Psychiatry 156: 675–82.

82. Oltedal L, Bartsch H, Sørhaug OJE, Kessler U, Abbott C, Dols A, et al. (2017): The Global ECT-MRI Research Collaboration (GEMRIC): Establishing a multi-site investigation of the neural mechanisms underlying response to electroconvulsive therapy. NeuroImage Clin 14: 422–432.

83. Price JL, Drevets WC (2010): Neurocircuitry of mood disorders. Neuropsychopharmacol Off Publ Am Coll Neuropsychopharmacol 35: 192–216.

84. Kahhale I, Buser NJ, Madan CR, Hanson JL (2023): Quantifying numerical and spatial reliability of hippocampal and amygdala subdivisions in FreeSurfer. Brain Inform 10: 9.

