## Supplemental for "Brain morphometry, stimulation charge, and seizure duration in electroconvulsive therapy"

### Supplemental Methods

**Data Selection.** Once combined, the original source datasets included 199 total participants with MRI, demographic, and clinical data. Of these, 4 did not receive right unilateral ECT, an additional 26 had missing or incomplete ECT treatment data, and a further 3 had stimulation charge values that were 3 standard deviations above or below the mean for each of the first three ECT sessions; data from these participants were omitted from further analysis. Data from the remaining n=166 were included in analyses as reflected in **Table 1**. Additional exclusions were made for single statistical tests based on MRI QC procedures per metric, or because ECT treatment data were missing from ECT1 or ECT2&3 (not both). Degrees of freedom are reported for each statistical analysis and reflect sample size (and regressors) used for each reported result.

**ECT Parameters: Methods and Rationale.** We chose stimulation dose in total charge as a single measure reflecting the amount of stimulation applied during treatment. If stimulation charge or one of its components was not recorded for a given ECT session, the following formula was used to calculate the missing value:

$$\text{Charge} = \text{amplitude} \times \text{pulse width} \times \text{frequency} \times \text{paired pulse duration}$$

The rationale for choosing stimulation charge was to mitigate differences in amplitude across cohorts (i.e., experimentally varied in UNM2 (1); 800mA for UNM1, UCLA1, UCLA2), as well as variations in pulse duration and/or frequency across patients, sessions, and cohorts (e.g., see Figure S1). Averaging charge over ECT sessions 2 and 3 also addresses any changes in these parameters across sessions. Though each stimulation parameter comprising charge is likely to have its own unique effects on (or interactions with) brain morphometry and function (2), future prospective studies could better examine the effects of each stimulation parameter (e.g., by independently modulating each parameter in animal models as in (3)).

Seizure duration was recorded at each ECT session using 2-channel EEG, and the current study uses this parameter as a measure of variability in seizure expression across study participants. Though it is possible to record other seizure characteristics using ECT devices, and qualitative measures of seizure expression are annotated, these parameters were not recorded as consistently and so were not chosen for analysis in the current study. For UCLA1 cohort, stimulus charge and seizure duration was available only for the first three ECT sessions and the final ECT session.

**MRI Acquisition & Preprocessing.** Baseline MRI data were collected before treatment using a 3T Siemens scanner at (or approximating) 1 $mm^3$ isovoxel resolution for each cohort; details and sequence parameters are described in source publications [e.g., UCLA1 (4), UCLA2 (5), UNM1 (6), UNM2 (7)]. The majority of volunteers had both T1- and T2-weighted MRI; both were used in analyses when available. If a T2-weighted image was not available, only the T1 was used (n=1 UCLA1, n=1 UNM1, n=11 UNM2).

Anatomical images were processed and morphological metrics were calculated using Freesurfer 7.2.0 (8–11). Subcortical and white matter volumes were extracted using the standard aseg, aparc.2009s, and wmparc atlases in Freesurfer's recon-all command (11), and add-on functions from Freesurfer calculated volumes for hippocampal subregions (12), amygdala nuclei (13), and thalamic nuclei (12). Cortical metrics from recon-all included thickness, surface area, and curvature using the aparc.2009s atlas (8).

Electric fields (E-fields) were estimated using SimNIBS (14,15). In brief, tissue segmentations derived from T1w and T2w images (gray matter, white matter, cerebrospinal fluid, skull, and scalp) were used to create tetrahedral volume conductor model or "head mesh" for each individual, and finite element models (FEMs) estimated unique patterns of current flow (E-field) between ECT electrodes in each individual head. Electrodes (5cm diameter circles) were positioned 2cm above the midpoint between right tragus and external canthus and 2cm to the right of the vertex (16). Standard recommended values for tissue conductivities were used (17). E-field magnitude (|E|, V/m) was calculated for current amplitude used during treatment, $|E|_{verum}$, and at the standard for ultra-brief pulse RUL ECT, 800mA (16), $|E|_{standard}$. Nifti $|E|_{verum}$ and $|E|_{standard}$ maps were registered to Freesurfer images in native space and parcellated using the same atlases as used for morphometric analyses. $|E|_{standard}$ was selected as a standard/uniform metric reflecting the ability of the electrical stimulus to penetrate the head during RUL ECT (regardless of stimulation amplitude used), because some UNM2 patients did not receive 800 mA stimulation. $|E|_{standard}$ is linearly related to a similar metric used by Lee et al. (18) and Deng, Abbott et al. (7,19), who calculated ratios between $|E|_{verum}$ and electrode or coil current, I. Because they are linearly related, our present statistical results would be identical for $|E|_{standard}$ and metrics used by these previous studies.

Each metric type (gray matter volume, white matter volume, cortical thickness, cortical area, cortical curvature, and |E|) was harmonized across cohorts with neuroCombat (20), applied separately for each metric type in R (https://www.r-project.org). NeuroCombat uses an empirical Bayesian approach originally developed to mitigate batch effects in genomics (21), and has been successfully applied to structural and functional MRI metrics in a variety of contexts (20,22–25).

**Statistics, Significance & Effect Size.** In main statistical analyses, metrics reflecting macro-anatomical structures were reported at a statistical criterion of uncorrected $p < 0.05$, and statistics for all other metrics were reported with correction for multiple comparisons using the false discovery rate (FDR) $q < 0.05$ (Benjamini-Hochberg method), applied separately for each model and metric type. For main analyses of |E|, we report results meeting both FDR $q < 0.05$ for $|E|_{verum}$ and uncorrected $p < 0.05$ for $|E|_{standard}$ (i.e., a conjunction of these two criteria), because stimulation amplitude contributes both to $|E|_{verum}$ and modestly to stimulation charge. Effect size is reported as partial $r^2$ (partial_r2 function, (26)), an estimate of the unique variance explained by each model term.

**Statistics, Follow-up Analyses.** Follow-up analyses were performed to aid in interpretation of the main analyses of stimulation charge, and to address potential cross-site differences in ECT administration (e.g., experimental manipulation of amplitude in the UNM2 cohort). Follow-up analyses were performed for each main analysis result for stimulation charge meeting statistical criteria as described above (for main analyses). First, separate statistical models replaced dependent variable (DV) charge with either number of pulses or amplitude. Independent variable of interest was MRI metric or $|E|_{standard}$, and regressors of no interest were age, sex, cohort, and amplitude (when pulse number was the DV) or pulse number (when amplitude was DV). Our hypothesis was that these models would be $p < 0.05$ for pulse number, but not amplitude. Datapoints with pulse width = 1.0 from UNM2 were excluded (n=12) (1). In a second set of follow-up analyses, statistical models were applied separately for UNM2 and UCLA cohorts. Models were constructed as in the main analysis, with charge as the dependent variable, MRI metric or |E| as the regressor of interest, and age and sex as regressors of no interest. Results for follow-up analyses are reported uncorrected $p < 0.05$.

Additional follow-up analyses determined whether regional metrics (morphometry or $|E|_{standard}$) identified in the main analyses differed between responders and nonresponders to RUL ECT. Here, response was defined as >50% improvement in Hamilton Depression Rating Scale (HDRS) score after RUL ECT, and nonresponse was defined as (1) < 50% improvement in HDRS score after RUL ECT, where only RUL treatments were given OR (2) transition to bitemporal (BT) ECT regardless of post-treatment HDRS score. Data for patients who did not complete ECT (Non-completers, NC) are displayed in figures, but were not included in these analyses. In these linear models, DV was MRI metric or $|E|_{standard}$, independent variable of interest was response (categorical factor), and variables of no interest were age, sex, and cohort. Results are reported uncorrected $p < 0.05$, and relationships amongst variables meeting this criterion were compared pairwise using Pearson's correlation coefficient. In this analysis, a negative correlation between cortical surface area near the right temporal electrode and $|E|_{standard}$ in anterior medial temporal lobe structures was identified. A mediation analysis explored this result further as described in the next section.

**Statistics, Mediation Analysis.** A mediation analysis probed whether $|E|_{standard}$ in anterior medial temporal lobe (AMTL) regions mediated (explained) the relationship between cortical surface area and antidepressant response to RUL ECT (mediation package in R (27)). Linear models were used to remain consistent with other statistics reported in this manuscript. For the full model, dependent variable was response (binary, numeric), with response defined as >50% improvement in HDRS score and nonresponse <50% improvement in HDRS score or transition to BT ECT. Independent variables were surface area of the anterior insula, $|E|_{standard}$ averaged across all AMTL regions identified in the follow-up analysis of response, as well as age, sex, and cohort. For the mediation model, dependent variable was mean AMTL $|E|_{standard}$ (mediator), and independent variables were anterior insula surface area (treatment), age, sex, and cohort. The mediate function was used, with confidence intervals estimated with nonparametric bootstrapping using the percentile method (5000 simulations). Surface area and $|E|_{standard}$ were z-normalized prior to this analysis.

### Supplemental Results

**ECT Parameters**. Because UNM2 cohort participants were randomly assigned to 600, 700, or 800mA stimulation amplitude, additional analyses assessed relationships between stimulation charge, amplitude, and other components of stimulation dose in each cohort separately. The rationale was to demonstrate that charge was not simply a recapitulation of amplitude. In these analyses, stimulation charge and amplitude were not strongly related in the UNM2 cohort ($r = -0.23$ in ECT1 and $r = -0.34$ in ECT2&3, $p > 0.05$ for both; **Figure S1**). Instead, stimulation charge was positively correlated with paired-pulse duration ($r = 0.395$ for ECT1; $r = 0.928$ for 2&3) and pulse width ($r = 0.725$ for ECT1; $r = 0.628$ for ECT2&3) in the UNM2 cohort ($p < 0.05$ for all; **Figure S1)**. This suggests that the higher range of stimulation charge in the UNM2 cohort was not explained well by amplitude, but instead reflects the combination of multiple stimulus parameters, just as in other cohorts.

Though charge (and number of pulses) during early sessions was marginally higher in participants who did not respond to RUL ECT (including transition to BT ECT), these differences appeared to be driven by outliers and were not considered further in the current manuscript (**Table S2; Figure S2**). Seizure duration did not differ in people who did not respond to RUL ECT (**Table S2; Figure S2**).

### Supplemental Discussion

#### Regarding Seizure Titration

During the first ECT session, each patient's seizure threshold is estimated, and electrical dose on subsequent sessions is increased in reference to this threshold estimate (e.g., 6x seizure threshold in RUL ECT). In the titration method, a low dose of stimulation is incrementally increased until a seizure of desired morphology occurs; however, seizure threshold can also be estimated, albeit crudely, using the patient's age, sex, and/or past history with ECT (28,29). In the present study, the range of stimulation charge was wider for UNM cohorts during the first session, reflecting the experimental dose titration procedure used in the UNM2 cohort (1). The UCLA cohorts had a narrower range of stimulation charge on first session, where seizure titration followed the recommended schedule (protocol) for the MECTA device (Spectrum Q 100 Joule Titration Table for Ultrabrief 0.3ms Pulsewidth, (30)). Here, 80% of UCLA patients had threshold at step #2 (i.e., 20Hz frequency, 0.3ms width, 2s duration; **Figure S1**). There are advantages and disadvantages to both approaches. Though seizure titration may provide a more accurate, individualized estimate of seizure threshold, excess stimulation may be a concern (e.g., if multiple rounds are needed to elicit seizure). Beginning titration with a higher dose more likely to elicit a seizure minimizes this risk; however, this method may over-estimate dose required for subsequent sessions (31), potentially increasing risk for cognitive or other side effects. Nevertheless, in the present study, stimulation dose at first session was much less than the dose used in subsequent sessions, and is therefore likely to be much nearer to a patient's seizure threshold, even if methods differed across sites.

#### Mid-Hippocampus and ECT2&3 Charge

We interpret correlation between mid-hippocampal volume and ECT2&3 charge as reflecting its role in seizure generalization, perhaps via connections with thalamus. However, other interpretations are possible. Plasticity in mid-hippocampus after ECT has been linked with antidepressant response (32,33), so it is possible that the relationship between the size of this region and stimulation charge also reflects the importance of this region to antidepressant response. However, pre-treatment mid-hippocampus volume did not differ in nonresponders to RUL ECT in the current study (though when nonresponders did not include people who transitioned to BT ECT, mid-hippocampal volume was modestly higher in people who completed but did not respond to RUL ECT, similar to (4)). Though not measured in the current study, post-ECT plasticity may be more relevant; Deng, Abbott, and colleagues reported that increased total hippocampal volume after ECT mediated the relationship between total hippocampal |E| and antidepressant response (7). However, it is also possible that the size/length of mid-hippocampus affects the position of anterior hippocampus and amygdala within the E-field distribution for RUL ECT, thus influencing susceptibility to therapeutic seizure and antidepressant response. Here, studies precisely measuring electrode position (or stimulation area) and resulting E field in relation to the position of AMTL structures in each patient could be informative.

#### |E| in Cortical White Matter

The primary effects of electrical stimulation in ECT and other brain stimulation technologies are generally assumed to occur in neuronal cell bodies and gray matter. However, other cells, including oligodendrocytes comprising white matter, astrocytes, interneurons, and even parts of the neurovascular unit (e.g., vascular smooth muscle) can all be influenced by electrical current (34–37). Indeed, this was the rationale for including both white and gray matter parcellations in our study. In addition, previous studies have also noted increased white matter integrity after ECT (38,39). Stimulation charge at both ECT1 and ECT2&3 correlated with |E| in white matter in right superior temporal cortex, as well as dorsal anterior cingulate and adjacent corpus callosum. Given that white matter is more conductive than gray matter, one possible interpretation of this finding is that electrical current (and thus seizure activity) may travel further from stimulating electrodes in patients with greater white matter |E|. For example, perhaps patients with more |E| reaching corpus callosum require less stimulation charge to elicit bilateral seizure activity at ECT1 (because the electrical stimulus is more likely to travel to the left hemisphere). Future studies incorporating EEG to measure seizure expression in right and left hemisphere in these patients could address this idea more directly.

#### ECT2&3 Seizure Duration and Subgenual Anterior and Posterior Cingulate Cortex (sgACC, PCC) Surface Area

Both the sgACC and PCC are implicated in self-referential processing and depressive neurobiology (40,41); however, it is unclear why surface area in these regions statistically predict seizure duration at supra-threshold stimulation in the current study. Though these metrics were not related to antidepressant response using our statistical criteria, pre-treatment surface area in sgACC was marginally smaller in responders to RUL ECT ($t_{117}$=1.93, p=0.06, partial $r^2$=0.03). In pairwise comparisons, sgACC surface area did not differ strongly between responders and nonresponders who continued RUL ECT ($t_{150}$=1.32, p=0.19, partial $r^2$=0.01) or transitioned to BT ECT ($t_{150}$=1.70, p=0.09, partial $r^2$=0.02). But, surface area for patients who did not complete ECT did appear to be more consistently larger compared with responders, though effect size was small ($t_{150}$=2.10, p=0.04, partial $r^2$=0.03). Given that patients with larger left sgACC tended to have longer seizures in this study, we speculate that these effects could be related to the size of nearby piriform cortex, which extends posteriorly from sgACC to entorhinal cortex and was not studied in this paper. Piriform cortex is a highly seizure-genic and understudied structure (42–44), and future studies directly measuring its contribution to ECT could be informative

**Regarding the Selection of Stimulation Charge in Main Analyses**

One of the goals of this study was to measure relationships between pre-treatment morphology and the amount of stimulation needed (or given) to elicit seizures during ECT. The main way we chose to measure the amount of stimulation given during treatment is stimulation charge (i.e., Total Charge in mC). Though stimulation charge is a summary measure combining multiple stimulation parameters, it has advantages. First, there is a history of using charge in previous ECT research, though its utility may be debated. Stimulation Charge is also a known/measured quantity (i.e., compared with |E|, which is estimated and not directly measured). Perhaps most importantly, using stimulation charge in this retrospective study may mitigate differences in stimulation parameters used across cohorts (e.g., experimental manipulation of amplitude in UNM2) and across patients and sessions (e.g., differences in pulse width in UNM2, or in other parameters for all patients). Systematic and independent experimental manipulation of each of these parameters would be the ideal way to assess their individual impacts; however, this is best suited to prospective trials, animal research, or even larger multi-cohort studies than the current study.

To complement our main analysis approach using stimulation charge, we also used follow-up analyses to address differences in amplitude across sites. These analyses confirmed that effects of charge were mostly consistent across sites, and were not driven by amplitude. Instead, these effects appeared to be better explained by pulse number (though stimulation charge was typically still superior to pulse number). Though it was possible to calculate stimulation time instead of pulse number, the great majority of patients received ultra brief pulse stimulation (0.3s; n=154), and so these follow-up analyses omitted the few UNM2 patients who received longer pulse width stimulation as part of their clinical trial protocol (1.0s; n=12) and pulse number was analyzed.

Estimated magnitude of electrical current, or |E|, was another way to study the amount of stimulation given during treatment. Though a potentially powerful tool, there are limitations to estimating E-fields. As mentioned above, |E| is an estimate, rather than a direct measure of the electrical current reaching different head and brain tissues. The accuracy of this estimate depends on the fidelity of tissue segmentation, how well the resistance of each patient's head tissues matches values assumed by the software, quality of the input MRIs, and other factors. In addition, both $|E|_{verum}$ and stimulation charge are calculated using stimulation amplitude; requiring a conjunction of $|E|_{verum}$ pFDR < 0.05 and $|E|_{standard}$ p < 0.05 for statistical significance attempted to mitigate this issue. Despite these limitations, |E| provides unique information about how much electric current reaches different parts of the brain across different patients and continues to be a useful tool in brain stimulation research.

**Supplemental References**


1. Abbott CC, Quinn D, Miller J, Ye E, Iqbal S, Lloyd M, *et al.* (2021): Electroconvulsive Therapy Pulse Amplitude and Clinical Outcomes. *Am J Geriatr Psychiatry* 29: 166–178.
2. Peterchev AV, Rosa MA, Deng Z-D, Prudic J, Lisanby SH (2010): Electroconvulsive therapy stimulus parameters: rethinking dosage. *J ECT* 26: 159–174.
3. Peterchev AV, Deng Z-D, Sikes-Keilp C, Feuer EC, Rosa MA, Lisanby SH (2024, September 30): Optimal Frequency for Seizure Induction with Electroconvulsive Therapy and Magnetic Seizure Therapy. bioRxiv, p 2024.09.28.615333.
4. Joshi SH, Espinoza RT, Pirnia T, Shi J, Wang Y, Ayers B, *et al.* (2015): Structural plasticity of the hippocampus and amygdala induced by electroconvulsive therapy in major depression. *Biol Psychiatry* 79: 282–92.
5. Tozzi L, Anene ET, Gotlib IH, Wintermark M, Kerr AB, Wu H, *et al.* (2021): Convergence, preliminary findings and future directions across the four human connectome projects investigating mood and anxiety disorders. *NeuroImage* 245: 118694.

6. Abbott CC, Jones T, Lemke NT, Gallegos P, McClintock SM, Mayer AR, *et al.* (2014): Hippocampal structural and functional changes associated with electroconvulsive therapy response. *Transl Psychiatry* 4: e483.
7. Deng Z-D, Argyelan M, Miller J, Quinn DK, Lloyd M, Jones TR, *et al.* (2022): Electroconvulsive therapy, electric field, neuroplasticity, and clinical outcomes. *Mol Psychiatry* 27: 1676–1682.
8. Destrieux C, Fischl B, Dale A, Halgren E (2010): Automatic parcellation of human cortical gyri and sulci using standard anatomical nomenclature. *NeuroImage* 53: 1–15.
9. Fischl B, Salat DH, Busa E, Albert M, Dieterich M, Haselgrove C, *et al.* (2002): Whole brain segmentation: automated labeling of neuroanatomical structures in the human brain. *Neuron* 33: 341–355.
10. Fischl B, Dale AM (2000): Measuring the thickness of the human cerebral cortex from magnetic resonance images. *Proc Natl Acad Sci* 97: 11050–11055.
11. Dale AM, Fischl B, Sereno MI (1999): Cortical surface-based analysis. I. Segmentation and surface reconstruction. *NeuroImage* 9: 179–194.
12. Iglesias JE, Augustinack JC, Nguyen K, Player CM, Player A, Wright M, *et al.* (2015): A computational atlas of the hippocampal formation using ex vivo, ultra-high resolution MRI: Application to adaptive segmentation of in vivo MRI. *NeuroImage* 115: 117–137.
13. Saygin ZM, Kliemann D, Iglesias JE, van der Kouwe AJW, Boyd E, Reuter M, *et al.* (2017): High-resolution magnetic resonance imaging reveals nuclei of the human amygdala: manual segmentation to automatic atlas. *NeuroImage* 155: 370–382.
14. Thielscher A, Antunes A, Saturnino GB (2015): Field modeling for transcranial magnetic stimulation: A useful tool to understand the physiological effects of TMS? *2015 37th Annual International Conference of the IEEE Engineering in Medicine and Biology Society (EMBC)* 222–225.
15. Saturnino GB, Puonti O, Nielsen JD, Antonenko D, Madsen KH, Thielscher A (2019): SimNIBS 2.1: A Comprehensive Pipeline for Individualized Electric Field Modelling for Transcranial Brain Stimulation. In: Makarov S, Horner M, Noetscher G, editors. *Brain and Human Body Modeling: Computational Human Modeling at EMBC 2018*. Cham (CH): Springer. Retrieved January 22, 2021, from http://www.ncbi.nlm.nih.gov/books/NBK549569/
16. Kellner CH, Knapp R, Husain MM, Rasmussen K, Sampson S, Cullum M, *et al.* (2010): Bifrontal, bitemporal and right unilateral electrode placement in ECT: randomised trial. *Br J Psychiatry J Ment Sci* 196: 226–34.
17. Burger HC, Milaan JB van (1943): Measurements of the specific Resistance of the human Body to direct Current. *Acta Med Scand* 114: 584–607.
18. Lee WH, Lisanby SH, Laine AF, Peterchev AV (2017): Minimum Electric Field Exposure for Seizure Induction with Electroconvulsive Therapy and Magnetic Seizure Therapy. *Neuropsychopharmacol Off Publ Am Coll Neuropsychopharmacol* 42: 1192–1200.
19. Abbott CC, Miller J, Farrar D, Argyelan M, Lloyd M, Squillaci T, *et al.* (2024): Amplitude-determined seizure-threshold, electric field modeling, and electroconvulsive therapy antidepressant and cognitive outcomes. *Neuropsychopharmacology* 49: 640–648.
20. Fortin J-P, Parker D, Tunç B, Watanabe T, Elliott MA, Ruparel K, *et al.* (2017): Harmonization of multi-site diffusion tensor imaging data. *NeuroImage* 161: 149–170.
21. Johnson WE, Li C, Rabinovic A (2007): Adjusting batch effects in microarray expression data using empirical Bayes methods. *Biostat Oxf Engl* 8: 118–127.
22. Fortin J-P, Cullen N, Sheline YI, Taylor WD, Aselcioglu I, Cook PA, *et al.* (2018): Harmonization of cortical thickness measurements across scanners and sites. *NeuroImage* 167: 104–120.
23. Yu M, Linn KA, Cook PA, Phillips ML, McInnis M, Fava M, *et al.* (2018): Statistical harmonization corrects site effects in functional connectivity measurements from multi-site fMRI data. *Hum Brain Mapp* 39: 4213–4227.
24. Radua J, Vieta E, Shinohara R, Kochunov P, Quidé Y, Green MJ, *et al.* (2020): Increased power by harmonizing structural MRI site differences with the ComBat batch adjustment method in ENIGMA. *NeuroImage* 218: 116956.
25. Cetin Karayumak S, Bouix S, Ning L, James A, Crow T, Shenton M, *et al.* (2019): Retrospective harmonization of multi-site diffusion MRI data acquired with different acquisition parameters. *NeuroImage* 184: 180–200.
26. Cinelli C, Hazlett C (2020): Making Sense of Sensitivity: Extending Omitted Variable Bias. *J R Stat Soc Ser B Stat Methodol* 82: 39–67.
27. Tingley D, Yamamoto T, Hirose K, Keele L, Imai K (2014): mediation: R Package for Causal Mediation Analysis. *J Stat Softw* 59: 1–38.
28. Francis-Taylor R, Ophel G, Martin D, Loo C (2020): The ictal EEG in ECT: A systematic review of the relationships between ictal features, ECT technique, seizure threshold and outcomes. *Brain Stimulat* 13: 1644–1654.
29. Kellner CH, Pritchett JT, Beale MD, Coffey CE (1997): *Handbook of ECT*. Washington, D.C.: American Psychiatric Press.
30. MECTA LLC (1997): spECTrum: MECTA Instruction Manual: 100 Joules Domestic. MECTA Corporation.
31. Luccarelli J, McCoy TH, Seiner SJ, Henry ME (2021): Total charge required to induce a seizure in a retrospective cohort of patients undergoing dose titration of right unilateral ultrabrief pulse ECT. *J ECT* 37: 40–45.
32. Leaver AM, Vasavada M, Kubicki A, Wade B, Loureiro J, Hellemann G, *et al.* (2020): Hippocampal subregions and networks linked with antidepressant response to electroconvulsive therapy. *Mol Psychiatry* 1–12.

33. Leaver AM, Vasavada M, Joshi SH, Wade B, Woods RP, Espinoza R, Narr KL (2019): Mechanisms of Antidepressant Response to Electroconvulsive Therapy Studied With Perfusion Magnetic Resonance Imaging. *Biol Psychiatry* 85: 466–476.
34. Bahr-Hosseini M, Bikson M (2021): Neurovascular-modulation: A review of primary vascular responses to transcranial electrical stimulation as a mechanism of action. *Brain Stimulat* 14: 837–847.
35. Wischnewski M, Alekseichuk I, Opitz A (2023): Neurocognitive, physiological, and biophysical effects of transcranial alternating current stimulation. *Trends Cogn Sci* 27: 189–205.
36. Mohan UR, Watrous AJ, Miller JF, Lega BC, Sperling MR, Worrell GA, *et al.* (2020): The effects of direct brain stimulation in humans depend on frequency, amplitude, and white-matter proximity. *Brain Stimulat* 13: 1183–1195.
37. Liu A, Vöröslakos M, Kronberg G, Henin S, Krause MR, Huang Y, *et al.* (2018): Immediate neurophysiological effects of transcranial electrical stimulation [no. 1]. *Nat Commun* 9: 5092.
38. Lyden H, Espinoza RT, Pirnia T, Clark K, Joshi SH, Leaver AM, *et al.* (2014): Electroconvulsive therapy mediates neuroplasticity of white matter microstructure in major depression. *Transl Psychiatry* 4: e380.
39. Repple J, Meinert S, Bollettini I, Grotegerd D, Redlich R, Zaremba D, *et al.* (2020): Influence of electroconvulsive therapy on white matter structure in a diffusion tensor imaging study. *Psychol Med* 50: 849–856.
40. Mayberg HS, Liotti M, Brannan SK, McGinnis S, Mahurin RK, Jerabek PA, *et al.* (1999): Reciprocal Limbic-Cortical Function and Negative Mood: Converging PET Findings in Depression and Normal Sadness. *Am J Psychiatry* 156: 675–82.
41. Spreng RN, Grady CL (2010): Patterns of Brain Activity Supporting Autobiographical Memory, Prospection, and Theory of Mind, and Their Relationship to the Default Mode Network. *J Cogn Neurosci* 22: 1112–1123.
42. Borger V, Schneider M, Taube J, Potthoff A-L, Keil VC, Hamed M, *et al.* (2021): Resection of piriform cortex predicts seizure freedom in temporal lobe epilepsy. *Ann Clin Transl Neurol* 8: 177–189.
43. Piredda S, Gale K (1985): A crucial epileptogenic site in the deep prepiriform cortex. *Nature* 317: 623–625.
44. Steinbart D, Yaakub SN, Steinbrenner M, Guldin LS, Holtkamp M, Keller SS, *et al.* (2023): Automatic and manual segmentation of the piriform cortex: Method development and validation in patients with temporal lobe epilepsy and Alzheimer's disease. *Hum Brain Mapp* 44: 3196–3209.

### Supplemental Tables

**Table S1, EEG Duration x Stimulation Charge controlling for age, sex, cohort**

| | | t | df | p | β | βse | partial $r^2$ |
|---|---|---|---|---|---|---|---|
| Seizure Duration, ECT1 | charge | -1.60 | 152 | 0.1111 | -0.29 | 0.18 | 0.017 |
| | age | -2.00 | 152 | 0.0476 | -0.40 | 0.20 | 0.026 |
| | sex | -1.89 | 152 | 0.0601 | -10.29 | 5.43 | 0.023 |
| | cohortUCLA2 | 0.53 | 152 | 0.5991 | 3.44 | 6.54 | 0.002 |
| | cohortUNM1 | -1.27 | 152 | 0.2059 | -17.85 | 14.05 | 0.011 |
| | cohortUNM2 | -0.63 | 152 | 0.5322 | -4.78 | 7.63 | 0.003 |
| Stimulation Charge, ECT1 | EEGduration | -1.60 | 152 | 0.1111 | -0.06 | 0.04 | 0.017 |
| | age | 1.39 | 152 | 0.1667 | 0.12 | 0.09 | 0.013 |
| | sex | -2.96 | 152 | 0.0036 | -6.92 | 2.34 | 0.054 |
| | cohortUCLA2 | 0.61 | 152 | 0.5407 | 1.75 | 2.86 | 0.002 |
| | cohortUNM1 | -0.33 | 152 | 0.7449 | -2.02 | 6.18 | 0.001 |
| | cohortUNM2 | 3.02 | 152 | 0.0030 | 9.81 | 3.25 | 0.057 |
| Seizure Duration, ECT2&3 | charge | -1.82 | 153 | 0.0714 | -0.04 | 0.02 | 0.021 |
| | age | -2.57 | 153 | 0.0112 | -0.30 | 0.12 | 0.041 |
| | sex | -1.14 | 153 | 0.2551 | -3.58 | 3.13 | 0.008 |
| | cohortUCLA2 | 1.04 | 153 | 0.3009 | 4.00 | 3.86 | 0.007 |
| | cohortUNM1 | -1.92 | 153 | 0.0569 | -15.82 | 8.25 | 0.023 |
| | cohortUNM2 | 0.12 | 153 | 0.9011 | 0.57 | 4.54 | 0.000 |
| Stimulation Charge, ECT2&3 | EEGduration | -1.82 | 153 | 0.0714 | -0.55 | 0.31 | 0.021 |
| | age | 1.54 | 153 | 0.1259 | 0.69 | 0.45 | 0.015 |
| | sex | -1.13 | 153 | 0.2615 | -13.48 | 11.96 | 0.008 |
| | cohortUCLA2 | 1.79 | 153 | 0.0752 | 26.17 | 14.61 | 0.021 |
| | cohortUNM1 | -0.86 | 153 | 0.3891 | -27.44 | 31.76 | 0.005 |
| | cohortUNM2 | 3.37 | 153 | 0.0009 | 56.38 | 16.71 | 0.069 |
| Seizure Duration, Sessions 2+ | charge | -3.97 | 159 | 0.0001 | -0.05 | 0.01 | 0.090 |
| | age | -2.22 | 159 | 0.0279 | -0.21 | 0.09 | 0.030 |
| | sex | -1.97 | 159 | 0.0508 | -4.93 | 2.50 | 0.024 |
| | cohortUCLA2 | -0.66 | 159 | 0.5086 | -2.09 | 3.16 | 0.003 |
| | cohortUNM1 | -2.35 | 159 | 0.0200 | -15.87 | 6.75 | 0.034 |
| | cohortUNM2 | -1.32 | 159 | 0.1903 | -5.01 | 3.81 | 0.011 |
| Stimulation Charge, Sessions 2+ | EEGduration | -3.97 | 159 | 0.0001 | -1.73 | 0.44 | 0.090 |
| | age | 0.99 | 159 | 0.3237 | 0.54 | 0.55 | 0.006 |
| | sex | -3.01 | 159 | 0.0030 | -42.76 | 14.18 | 0.054 |
| | cohortUCLA2 | 1.62 | 159 | 0.1081 | 29.19 | 18.06 | 0.016 |
| | cohortUNM1 | -0.11 | 159 | 0.9130 | -4.32 | 39.52 | <0.001 |
| | cohortUNM2 | 4.62 | 159 | <0.0001 | 95.55 | 20.67 | 0.118 |

**Table S2, Responder vs. Nonresponder differences controlling for age, sex, cohort**

| | t | df | p | β | βse | partial_r2 |
|---|---|---|---|---|---|---|
| ECT1 Charge | 1.86 | 116 | 0.065 | 0.006 | 0.003 | 0.029 |
| ECT1 EEGduration | 1.40 | 116 | 0.164 | 0.002 | 0.001 | 0.017 |
| ECT1 # of Pulses | 2.49 | 113 | 0.014 | 0.004 | 0.002 | 0.052 |
| ECT2&3 Charge | 1.75 | 115 | 0.083 | 0.001 | 0.001 | 0.026 |
| ECT2&3 EEGduration | 0.38 | 115 | 0.706 | 0.001 | 0.002 | 0.001 |
| ECT2&3 # of Pulses | 2.03 | 114 | 0.045 | 0.001 | 0.000 | 0.035 |

**Table S3. Macro-morphometry**

| Metric, Session | Measure | t | df | p | β | βse | partial $r^2$ |
|---|---|---|---|---|---|---|---|
| Seizure Duration, ECT1 | Total Intracranial Volume | 2.20 | 151 | 0.03 | <0.0001 | <0.0001 | 0.031 |
| | L Cortical GMV | 2.50 | 151 | 0.01 | 0.0004 | 0.0002 | 0.040 |
| | R Cortical GMV | 2.77 | 151 | 0.006 | 0.0004 | 0.0002 | 0.048 |
| | Subcortical GMV | 0.82 | 151 | 0.42 | 0.0005 | 0.0006 | 0.004 |
| | L Cerebral WMV | 0.32 | 151 | 0.75 | <0.0001 | 0.0001 | 0.001 |
| | R Cerebral WMV | 0.53 | 151 | 0.60 | 0.0001 | 0.0001 | 0.002 |
| | CSF | -1.02 | 151 | 0.31 | -0.0105 | 0.0102 | 0.007 |
| Stimulation Charge, ECT1 | Total Intracranial Volume | 2.38 | 151 | 0.02 | <0.0001 | <0.0001 | 0.036 |
| | L Cortical GMV | 2.62 | 151 | 0.01 | 0.0002 | 0.0001 | 0.044 |
| | R Cortical GMV | 2.77 | 151 | 0.006 | 0.0002 | 0.0001 | 0.048 |
| | Subcortical GMV | 1.64 | 151 | 0.10 | 0.0004 | 0.0003 | 0.017 |
| | L Cerebral WMV | 0.80 | 151 | 0.43 | <0.0001 | <0.0001 | 0.004 |
| | R Cerebral WMV | 0.81 | 151 | 0.42 | <0.0001 | <0.0001 | 0.004 |
| | CSF | 0.74 | 151 | 0.46 | 0.0033 | 0.0045 | 0.004 |
| Seizure Duration, ECT2&3 | Total Intracranial Volume | 1.79 | 152 | 0.07 | <0.0001 | <0.0001 | 0.021 |
| | L Cortical GMV | 3.01 | 152 | 0.003 | 0.0003 | 0.0001 | 0.056 |
| | R Cortical GMV | 3.13 | 152 | 0.002 | 0.0003 | 0.0001 | 0.061 |
| | Subcortical GMV | 1.12 | 152 | 0.27 | 0.0004 | 0.0003 | 0.008 |
| | L Cerebral WMV | 0.21 | 152 | 0.83 | <0.0001 | 0.0001 | 0.000 |
| | R Cerebral WMV | 0.45 | 152 | 0.66 | <0.0001 | 0.0001 | 0.001 |
| | CSF | -0.33 | 152 | 0.74 | -0.0020 | 0.0060 | 0.001 |
| Stimulation Charge, ECT2&3 | Total Intracranial Volume | 1.64 | 152 | 0.10 | 0.0001 | <0.0001 | 0.017 |
| | L Cortical GMV | 2.37 | 152 | 0.02 | 0.0008 | 0.0003 | 0.035 |
| | R Cortical GMV | 2.42 | 152 | 0.02 | 0.0008 | 0.0003 | 0.037 |
| | Subcortical GMV | 2.48 | 152 | 0.01 | 0.0031 | 0.0013 | 0.039 |
| | L Cerebral WMV | 0.65 | 152 | 0.52 | 0.0001 | 0.0002 | 0.003 |
| | R Cerebral WMV | 0.60 | 152 | 0.55 | 0.0001 | 0.0002 | 0.002 |
| | CSF | 0.68 | 152 | 0.50 | 0.0156 | 0.0230 | 0.003 |

**Table S4. Regional morphometry and pulse number**

| | | Number of Pulses | | | | | | Amplitude | | | | | |
|---|---|---|---|---|---|---|---|---|---|---|---|---|---|
| Session | Region, Measure | t | df | p | β | βse | partial $r^2$ | t | df | p | β | βse | partial $r^2$ |
| ECT1 | R Inferior Frontal Gyrus Opercular Area, Mean Curv | 2.06 | 138 | 0.0411 | 507.2 | 246.0 | 0.030 | -0.10 | 140 | 0.9198 | -0.052 | 0.512 | 0.000 |
| | R Inferior Temporal Sulcus, SA | 3.57 | 138 | 0.0005 | 0.050 | 0.014 | 0.084 | -1.55 | 140 | 0.1232 | 0.000 | 0.000 | 0.017 |
| | L Paracentral Gyrus and Sulcus, SA | 2.62 | 138 | 0.0098 | 0.051 | 0.020 | 0.047 | 0.61 | 140 | 0.5417 | 0.000 | 0.000 | 0.003 |
| | R Circular Sulcus, Superior Insula, SA | 1.07 | 138 | 0.2857 | 0.023 | 0.022 | 0.008 | 0.48 | 140 | 0.6326 | 0.000 | 0.000 | 0.002 |
| | R Posterior Lateral Fissure, SA | 0.78 | 138 | 0.4369 | 0.019 | 0.024 | 0.004 | 0.53 | 140 | 0.5971 | 0.000 | 0.000 | 0.002 |
| | L Superior Frontal Gyrus, SA | 1.95 | 138 | 0.0528 | 0.007 | 0.004 | 0.027 | -1.02 | 140 | 0.3101 | 0.000 | 0.000 | 0.007 |
| | R Inferior Frontal Gyrus Opercular Area, SA | 1.40 | 138 | 0.1641 | 0.023 | 0.016 | 0.014 | -0.42 | 140 | 0.6758 | 0.000 | 0.000 | 0.001 |
| ECT2&3 | R Short Insular Gyrus, Mean Curv | 2.00 | 139 | 0.0469 | 1446.8 | 721.6 | 0.028 | 1.20 | 139 | 0.2338 | 0.344 | 0.288 | 0.010 |
| | L Thalamus, LP nucleus, Vol | 2.07 | 139 | 0.0406 | 1.106 | 0.535 | 0.030 | -0.39 | 139 | 0.6937 | 0.000 | 0.000 | 0.001 |
| | R Hippocampus Body, Dentate Gyrus et al., Vol | 3.17 | 125 | 0.0019 | 2.186 | 0.690 | 0.074 | 0.08 | 125 | 0.9378 | 0.000 | 0.000 | 0.000 |
| | R Thalamus, AV nucleus, Vol | 1.16 | 139 | 0.2471 | 0.628 | 0.541 | 0.010 | -0.64 | 139 | 0.5231 | 0.000 | 0.000 | 0.003 |
| | R Hippocampus Body CA4 region, Vol | 3.13 | 125 | 0.0022 | 2.513 | 0.804 | 0.073 | 0.41 | 125 | 0.6820 | 0.000 | 0.000 | 0.001 |
| | R Thalamus, LP nucleus, Vol | 1.82 | 139 | 0.0702 | 0.998 | 0.547 | 0.023 | -0.38 | 139 | 0.7060 | 0.000 | 0.000 | 0.001 |
| | L Thalamus, Inferior Pulvinar nucleus, Vol | 1.70 | 139 | 0.0922 | 0.616 | 0.363 | 0.020 | 0.04 | 139 | 0.9687 | 0.000 | 0.000 | 0.000 |
| | R Thalamus, VA nulceus, Vol | 2.09 | 139 | 0.0380 | 0.475 | 0.227 | 0.031 | 0.46 | 139 | 0.6457 | 0.000 | 0.000 | 0.002 |
| | R Thalamus, Total Vol | 1.77 | 139 | 0.0794 | 0.027 | 0.015 | 0.022 | -0.70 | 139 | 0.4842 | 0.000 | 0.000 | 0.004 |
| | R Thalamus CeM nulceus, Vol | 1.58 | 139 | 0.1157 | 1.631 | 1.030 | 0.018 | 0.58 | 139 | 0.5630 | 0.000 | 0.000 | 0.002 |

**Table S5. Regional morphometry and stimulation charge, separate cohorts**

| | | UNM2 Only | | | | | | UCLA1 & UCLA2 | | | | | |
|---|---|---|---|---|---|---|---|---|---|---|---|---|---|
| Session | Region, Measure | t | df | p | β | βse | partial $r^2$ | t | df | p | β | βse | partial $r^2$ |
| 1 | R Inferior Frontal Gyrus (Operculum), Mean Curv | 3.87 | 51 | 0.0003 | 927.34 | 239.5 | 0.227 | 1.77 | 92 | 0.0799 | 192.89 | 108.9 | 0.033 |
| | R Inferior Temporal Sulcus, SA | 2.62 | 51 | 0.0114 | 0.041 | 0.016 | 0.119 | 3.25 | 92 | 0.0016 | 0.020 | 0.006 | 0.103 |
| | L Paracentral Gyrus and Sulcus, SA | 3.25 | 51 | 0.0021 | 0.066 | 0.020 | 0.171 | 1.57 | 92 | 0.1189 | 0.015 | 0.009 | 0.026 |
| | R Circular Sulcus, Superior Insula, SA | 3.62 | 51 | 0.0007 | 0.088 | 0.024 | 0.205 | 1.55 | 92 | 0.1237 | 0.015 | 0.010 | 0.026 |
| | R Posterior Lateral Fissure, SA | 3.01 | 51 | 0.0041 | 0.078 | 0.026 | 0.151 | 1.50 | 92 | 0.1366 | 0.016 | 0.011 | 0.024 |
| | L Superior Frontal Gyrus, SA | 3.06 | 51 | 0.0035 | 0.012 | 0.004 | 0.155 | 1.66 | 92 | 0.0997 | 0.003 | 0.002 | 0.029 |
| | R Inferior Frontal Gyrus Opercular Area, SA | 3.33 | 51 | 0.0016 | 0.064 | 0.019 | 0.178 | 1.13 | 92 | 0.2617 | 0.008 | 0.007 | 0.014 |
| 2&3 | R Short Insular Gyrus, Mean Curv | 2.85 | 51 | 0.0062 | 2270.9 | 795.9 | 0.138 | 2.51 | 93 | 0.0138 | 706.2 | 281.4 | 0.063 |
| | L Thalamus, LP nucleus, Vol | 2.35 | 51 | 0.0225 | 1.367 | 0.581 | 0.098 | 2.24 | 93 | 0.0273 | 0.529 | 0.236 | 0.051 |
| | R Hippocampus Body, Dentate Gyrus et al., Vol | 2.53 | 40 | 0.0155 | 2.622 | 1.037 | 0.138 | 2.09 | 91 | 0.0391 | 0.594 | 0.284 | 0.046 |
| | R Thalamus, AV nucleus, Vol | 3.68 | 51 | 0.0006 | 2.161 | 0.588 | 0.210 | 1.18 | 93 | 0.2425 | 0.255 | 0.217 | 0.015 |
| | R Hippocampus Body CA4 region, Vol | 2.31 | 40 | 0.0260 | 2.702 | 1.168 | 0.118 | 2.09 | 91 | 0.0393 | 0.695 | 0.332 | 0.046 |
| | R Thalamus, LP nucleus, Vol | 2.56 | 51 | 0.0136 | 1.594 | 0.623 | 0.114 | 1.34 | 93 | 0.1848 | 0.310 | 0.232 | 0.019 |
| | L Thalamus, Inferior Pulvinar nucleus, Vol | 2.49 | 51 | 0.0161 | 1.074 | 0.432 | 0.108 | 2.77 | 93 | 0.0068 | 0.383 | 0.139 | 0.076 |
| | R Thalamus, VA nucleus, Vol | 2.48 | 51 | 0.0166 | 0.753 | 0.304 | 0.107 | 2.88 | 93 | 0.0049 | 0.250 | 0.087 | 0.082 |
| | R Thalamus, Total Vol | 2.97 | 51 | 0.0046 | 0.051 | 0.017 | 0.147 | 1.71 | 93 | 0.0914 | 0.011 | 0.006 | 0.030 |
| | R Thalamus CeM nucleus, Vol | 2.59 | 51 | 0.0124 | 3.136 | 1.209 | 0.117 | 1.81 | 93 | 0.0729 | 0.769 | 0.424 | 0.034 |

**Table S6. Estimated $|E|_{standard}$ and Pulse Number**

| | | Number of Pulses | | | | | | Amplitude | | | | | |
|---|---|---|---|---|---|---|---|---|---|---|---|---|---|
| Session | Region, Metric | t | df | p | β | βse | partial $r^2$ | t | df | p | β | βse | partial $r^2$ |
| 1 | R Superior Temporal WMV | -2.38 | 137 | 0.019 | -0.16 | 0.07 | 0.040 | -0.76 | 137 | 0.450 | 0.00 | 0.00 | 0.004 |
| 1 | R Inferior Temporal WMV | -2.29 | 137 | 0.024 | -0.20 | 0.09 | 0.037 | -0.28 | 137 | 0.781 | 0.00 | 0.00 | 0.001 |
| 1 | R Caudal Anterior Cingulate WMV | -2.87 | 137 | 0.005 | -0.25 | 0.09 | 0.057 | -0.57 | 137 | 0.568 | 0.00 | 0.00 | 0.002 |
| 1 | R Transverse Temporal WMV | -1.96 | 137 | 0.052 | -0.18 | 0.09 | 0.027 | -0.88 | 137 | 0.381 | 0.00 | 0.00 | 0.006 |
| 2&3 | R Amygdala, CAT volume | -1.46 | 124 | 0.146 | -0.31 | 0.21 | 0.017 | -0.32 | 124 | 0.749 | 0.00 | 0.00 | 0.001 |
| 2&3 | R Hippocampus Head, HATA volume | -1.78 | 124 | 0.077 | -0.35 | 0.19 | 0.025 | -0.40 | 124 | 0.687 | 0.00 | 0.00 | 0.001 |
| 2&3 | R Amygdala, Total volume | -1.84 | 137 | 0.069 | -0.31 | 0.17 | 0.024 | 0.01 | 137 | 0.993 | 0.00 | 0.00 | 0.000 |
| 2&3 | R Superior Temporal WMV | -2.38 | 137 | 0.019 | -0.16 | 0.07 | 0.040 | -0.76 | 137 | 0.450 | 0.00 | 0.00 | 0.004 |
| 2&3 | R Amygdala, AB Volume | -1.11 | 124 | 0.268 | -0.24 | 0.21 | 0.010 | -0.05 | 124 | 0.959 | 0.00 | 0.00 | 0.000 |
| 2&3 | R Hippocampus, Parasubiculum Volume | -1.67 | 124 | 0.097 | -0.36 | 0.22 | 0.022 | 0.08 | 124 | 0.939 | 0.00 | 0.00 | 0.000 |
| 2&3 | R Amygdala, Basal Volume | -1.35 | 124 | 0.178 | -0.27 | 0.20 | 0.015 | 0.18 | 124 | 0.854 | 0.00 | 0.00 | 0.000 |
| 2&3 | R Transverse Temporal WMV | -1.96 | 137 | 0.052 | -0.18 | 0.09 | 0.027 | -0.88 | 137 | 0.381 | 0.00 | 0.00 | 0.006 |
| 2&3 | R Putamen Volume | -1.81 | 137 | 0.072 | -0.27 | 0.15 | 0.023 | 0.20 | 137 | 0.845 | 0.00 | 0.00 | 0.000 |
| 2&3 | R Hippocampus, Total Volume | -1.74 | 137 | 0.085 | -0.29 | 0.17 | 0.022 | -0.02 | 137 | 0.985 | 0.00 | 0.00 | 0.000 |
| 2&3 | R Lateral Orbitofrontal WMV | -1.82 | 137 | 0.072 | -0.18 | 0.10 | 0.023 | 0.32 | 137 | 0.753 | 0.00 | 0.00 | 0.001 |
| 2&3 | R Hippocampus Head, Dentate et al. Volume | -1.73 | 124 | 0.085 | -0.33 | 0.19 | 0.024 | 0.16 | 124 | 0.873 | 0.00 | 0.00 | 0.000 |
| 2&3 | R Inf Lateral Ventricle Volume | -1.31 | 137 | 0.192 | -0.16 | 0.12 | 0.012 | -0.02 | 137 | 0.981 | 0.00 | 0.00 | 0.000 |
| 2&3 | R Globus Pallidus Volume | -1.78 | 137 | 0.077 | -0.23 | 0.13 | 0.023 | 0.22 | 137 | 0.828 | 0.00 | 0.00 | 0.000 |
| 2&3 | R Hippocampus Head, CA3 Volume | -1.68 | 124 | 0.095 | -0.31 | 0.19 | 0.022 | -0.09 | 124 | 0.930 | 0.00 | 0.00 | 0.000 |
| 2&3 | R Inferior Temporal WMV | -2.29 | 137 | 0.024 | -0.20 | 0.09 | 0.037 | -0.28 | 137 | 0.781 | 0.00 | 0.00 | 0.001 |
| 2&3 | R Caudal Anterior Cingulate WMV | -2.87 | 137 | 0.005 | -0.25 | 0.09 | 0.057 | -0.57 | 137 | 0.568 | 0.00 | 0.00 | 0.002 |
| 2&3 | R Amygdala, Paralaminar Volume | -1.54 | 124 | 0.127 | -0.30 | 0.19 | 0.019 | 0.03 | 124 | 0.977 | 0.00 | 0.00 | 0.000 |
| 2&3 | R Insula, WMV | -1.82 | 137 | 0.071 | -0.19 | 0.10 | 0.024 | 0.27 | 137 | 0.790 | 0.00 | 0.00 | 0.001 |
| 2&3 | R Inferior Frontal (Pars Triangularis) WMV | -2.11 | 137 | 0.037 | -0.17 | 0.08 | 0.031 | -0.05 | 137 | 0.957 | 0.00 | 0.00 | 0.000 |
| 2&3 | R Entorhinal WMV | -1.77 | 137 | 0.078 | -0.20 | 0.11 | 0.022 | -0.15 | 137 | 0.878 | 0.00 | 0.00 | 0.000 |
| 2&3 | R Hippocampus Head, CA4 Volume | -1.59 | 124 | 0.115 | -0.29 | 0.18 | 0.020 | 0.45 | 124 | 0.650 | 0.00 | 0.00 | 0.002 |
| 2&3 | R Parahippocampal WMV | -1.65 | 137 | 0.100 | -0.25 | 0.15 | 0.020 | -0.18 | 137 | 0.854 | 0.00 | 0.00 | 0.000 |
| 2&3 | R Inferior Frontal (Pars Orbitalis) WMV | -2.08 | 137 | 0.039 | -0.18 | 0.09 | 0.031 | 0.25 | 137 | 0.806 | 0.00 | 0.00 | 0.000 |

| | | | | | | | | | | | | | |
|---|---|---|---|---|---|---|---|---|---|---|---|---|---|
| 2&3 | R Rostral Middle Frontal WMV | -1.65 | 137 | 0.100 | -0.17 | 0.10 | 0.020 | 0.40 | 137 | 0.687 | 0.00 | 0.00 | 0.001 |
| 2&3 | R Middle Temporal WMV | -2.11 | 137 | 0.037 | -0.16 | 0.08 | 0.031 | -0.25 | 137 | 0.801 | 0.00 | 0.00 | 0.000 |
| 2&3 | R Hippocampus Head, CA1 Volume | -1.45 | 124 | 0.150 | -0.27 | 0.18 | 0.017 | 0.01 | 124 | 0.995 | 0.00 | 0.00 | 0.000 |
| 2&3 | R Fusiform WMV | -2.25 | 137 | 0.026 | -0.31 | 0.14 | 0.036 | -0.72 | 137 | 0.473 | 0.00 | 0.00 | 0.004 |
| 2&3 | R Hippocampus Head, Molecular Layer Volume | -1.58 | 124 | 0.116 | -0.28 | 0.17 | 0.020 | 0.27 | 124 | 0.791 | 0.00 | 0.00 | 0.001 |
| 2&3 | R Thalamus, Lateral Geniculate Nucleus Volume | -1.11 | 137 | 0.268 | -0.15 | 0.13 | 0.009 | 0.31 | 137 | 0.757 | 0.00 | 0.00 | 0.001 |
| 2&3 | Pons | -1.42 | 137 | 0.159 | -0.36 | 0.25 | 0.014 | -0.23 | 137 | 0.820 | 0.00 | 0.00 | 0.000 |
| 2&3 | R Inferior Frontal (Pars Opercularis) | -1.36 | 137 | 0.175 | -0.11 | 0.08 | 0.013 | 0.49 | 137 | 0.623 | 0.00 | 0.00 | 0.002 |

**Table S7. Estimated $|E|_{standard}$ and charge, separate cohorts**

| | | UNM2 | | | | | | UCLA | | | | | |
|---|---|---|---|---|---|---|---|---|---|---|---|---|---|
| Session | Region, Metric | t | df | p | β | βse | partial $r^2$ | t | df | p | β | βse | partial $r^2$ |
| 1 | R Superior Temporal WMV | -1.02 | 50 | 0.311 | -0.11 | 0.11 | 0.021 | -3.05 | 90 | 0.003 | -0.10 | 0.03 | 0.094 |
| 1 | R Inferior Temporal WMV | -0.69 | 50 | 0.492 | -0.10 | 0.14 | 0.010 | -2.87 | 90 | 0.005 | -0.13 | 0.04 | 0.084 |
| 1 | R Caudal Anterior Cingulate WMV | 0.18 | 50 | 0.860 | 0.02 | 0.13 | 0.001 | -4.09 | 90 | 0.000 | -0.16 | 0.04 | 0.157 |
| 1 | R Transverse Temporal WMV | -0.45 | 50 | 0.651 | -0.07 | 0.15 | 0.004 | -3.08 | 90 | 0.003 | -0.14 | 0.04 | 0.095 |
| 2&3 | R Amygdala, CAT volume | -1.71 | 39 | 0.095 | -2.66 | 1.55 | 0.070 | -2.27 | 93 | 0.025 | -0.93 | 0.41 | 0.053 |
| 2&3 | R Hippocampus Head, HATA volume | -1.38 | 39 | 0.175 | -1.90 | 1.37 | 0.047 | -2.75 | 93 | 0.007 | -1.04 | 0.38 | 0.075 |
| 2&3 | R Amygdala, Total volume | -1.92 | 50 | 0.061 | -2.78 | 1.45 | 0.069 | -2.08 | 94 | 0.040 | -0.77 | 0.37 | 0.044 |
| 2&3 | R Superior Temporal WMV | -1.80 | 50 | 0.078 | -1.05 | 0.58 | 0.061 | -2.05 | 94 | 0.043 | -0.29 | 0.14 | 0.043 |
| 2&3 | R Amygdala, AB Volume | -1.53 | 39 | 0.135 | -2.65 | 1.74 | 0.056 | -2.33 | 93 | 0.022 | -0.99 | 0.42 | 0.055 |
| 2&3 | R Hippocampus, Parasubiculum Volume | -2.52 | 39 | 0.016 | -4.89 | 1.94 | 0.140 | -1.21 | 93 | 0.231 | -0.54 | 0.45 | 0.015 |
| 2&3 | R Amygdala, Basal Volume | -1.95 | 39 | 0.059 | -3.77 | 1.93 | 0.089 | -1.80 | 93 | 0.074 | -0.72 | 0.40 | 0.034 |
| 2&3 | R Transverse Temporal WMV | -1.00 | 50 | 0.321 | -0.80 | 0.80 | 0.020 | -2.63 | 94 | 0.010 | -0.53 | 0.20 | 0.069 |
| 2&3 | R Putamen Volume | -1.08 | 50 | 0.285 | -1.45 | 1.34 | 0.023 | -2.68 | 94 | 0.009 | -0.84 | 0.31 | 0.071 |
| 2&3 | R Hippocampus, Total Volume | -1.42 | 50 | 0.163 | -2.15 | 1.52 | 0.039 | -2.01 | 94 | 0.048 | -0.74 | 0.37 | 0.041 |
| 2&3 | R Lateral Orbitofrontal WMV | -1.15 | 50 | 0.255 | -0.91 | 0.79 | 0.026 | -2.29 | 94 | 0.025 | -0.51 | 0.23 | 0.053 |
| 2&3 | R Hippocampus Head, Dentate et al. Volume | -1.94 | 39 | 0.060 | -3.61 | 1.86 | 0.088 | -1.77 | 93 | 0.080 | -0.68 | 0.38 | 0.033 |
| 2&3 | R Inf Lateral Ventricle Volume | -1.17 | 50 | 0.249 | -1.17 | 1.00 | 0.027 | -2.03 | 94 | 0.046 | -0.56 | 0.27 | 0.042 |
| 2&3 | R Globus Pallidus Volume | -0.68 | 50 | 0.502 | -0.77 | 1.14 | 0.009 | -3.01 | 94 | 0.003 | -0.81 | 0.27 | 0.088 |
| 2&3 | R Hippocampus Head, CA3 Volume | -1.72 | 39 | 0.093 | -2.97 | 1.72 | 0.071 | -1.92 | 93 | 0.058 | -0.74 | 0.39 | 0.038 |
| 2&3 | R Inferior Temporal WMV | -1.37 | 50 | 0.178 | -1.06 | 0.77 | 0.036 | -2.24 | 94 | 0.027 | -0.43 | 0.19 | 0.051 |
| 2&3 | R Caudal ACC WMV | -0.62 | 50 | 0.537 | -0.43 | 0.69 | 0.008 | -3.85 | 94 | 0.000 | -0.71 | 0.18 | 0.136 |
| 2&3 | R Amygdala, Paralaminar Volume | -1.89 | 39 | 0.067 | -3.41 | 1.81 | 0.084 | -1.60 | 93 | 0.113 | -0.63 | 0.40 | 0.027 |
| 2&3 | R Insula, WMV | -0.74 | 50 | 0.463 | -0.69 | 0.94 | 0.011 | -2.52 | 94 | 0.013 | -0.56 | 0.22 | 0.063 |
| 2&3 | R Inferior Frontal (Pars Triangularis) WMV | -0.75 | 50 | 0.455 | -0.53 | 0.70 | 0.011 | -2.89 | 94 | 0.005 | -0.52 | 0.18 | 0.082 |
| 2&3 | R Entorhinal WMV | -1.05 | 50 | 0.298 | -0.98 | 0.94 | 0.022 | -1.94 | 94 | 0.055 | -0.47 | 0.24 | 0.039 |
| 2&3 | R Hippocampus Head, CA4 Volume | -1.93 | 39 | 0.061 | -3.50 | 1.82 | 0.087 | -1.45 | 93 | 0.151 | -0.54 | 0.38 | 0.022 |
| 2&3 | R Parahippocampal WMV | -0.84 | 50 | 0.403 | -1.19 | 1.42 | 0.014 | -2.54 | 94 | 0.013 | -0.86 | 0.34 | 0.064 |

| | | | | | | | | | | | | | |
|---|---|---|---|---|---|---|---|---|---|---|---|---|---|
| 2&3 | R Inferior Frontal (Pars Orbitalis) WMV | -0.51 | 50 | 0.614 | -0.36 | 0.71 | 0.005 | -2.96 | 94 | 0.004 | -0.56 | 0.19 | 0.085 |
| 2&3 | R Rostral Middle Frontal WMV | -0.36 | 50 | 0.722 | -0.28 | 0.78 | 0.003 | -2.93 | 94 | 0.004 | -0.65 | 0.22 | 0.083 |
| 2&3 | R Middle Temporal WMV | -1.09 | 50 | 0.281 | -0.74 | 0.68 | 0.023 | -1.98 | 94 | 0.051 | -0.33 | 0.17 | 0.040 |
| 2&3 | R Hippocampus Head, CA1 Volume | -1.59 | 39 | 0.121 | -3.07 | 1.93 | 0.061 | -1.72 | 93 | 0.089 | -0.63 | 0.36 | 0.031 |
| 2&3 | R Fusiform WMV | -0.52 | 50 | 0.605 | -0.67 | 1.29 | 0.005 | -2.35 | 94 | 0.021 | -0.72 | 0.31 | 0.055 |
| 2&3 | R Hippocampus Head, Molecular Layer Volume | -1.58 | 39 | 0.121 | -2.86 | 1.80 | 0.060 | -1.58 | 93 | 0.117 | -0.56 | 0.35 | 0.026 |
| 2&3 | R Thalamus, Lateral Geniculate Nucleus Volume | -0.32 | 50 | 0.748 | -0.36 | 1.10 | 0.002 | -2.60 | 94 | 0.011 | -0.77 | 0.29 | 0.067 |
| 2&3 | Pons | -0.70 | 50 | 0.490 | -1.57 | 2.26 | 0.010 | -2.32 | 94 | 0.023 | -1.24 | 0.53 | 0.054 |
| 2&3 | R Inferior Frontal (Pars Opercularis) | -0.52 | 50 | 0.603 | -0.35 | 0.68 | 0.005 | -2.04 | 94 | 0.045 | -0.34 | 0.17 | 0.042 |

**Table S8. Differences in responders vs. nonresponders to RUL ECT p < 0.05**

| | | t | df | p | β | βse | partial $r^2$ |
|---|---|---|---|---|---|---|---|
| RUL R vs. RUL NR | R Inferior Frontal G Opercular Area, SA | 1.98 | 150 | 0.049 | 68.00 | 34.27 | 0.03 |
| | R Circular S, Superior Insula, SA | 1.50 | 150 | 0.136 | 38.346 | 25.600 | 0.015 |
| | R Inferior Frontal G Opercular Area, Mean Curv | 1.44 | 150 | 0.152 | 0.003 | 0.002 | 0.014 |
| | R Amygdala, Paralaminar Volume | -2.34 | 136 | 0.020 | -6.583 | 2.807 | 0.039 |
| | R Hippocampus, Parasubiculum Volume | -2.08 | 136 | 0.039 | -5.244 | 2.519 | 0.031 |
| | R Hippocampus Head, Molecular Layer Volume | -1.98 | 136 | 0.049 | -6.075 | 3.065 | 0.028 |
| | R Amygdala, Basal Volume | -2.04 | 136 | 0.044 | -5.626 | 2.761 | 0.030 |
| | R Hippocampus Head, CA1 Volume | -1.94 | 136 | 0.055 | -5.713 | 2.947 | 0.027 |
| | R Hippocampus Head, CA4 Volume | -1.56 | 136 | 0.122 | -4.565 | 2.935 | 0.017 |
| | R Hippocampus Head, Dentate et al. Volume | -1.54 | 136 | 0.126 | -4.400 | 2.854 | 0.017 |
| | R Amygdala, AB Volume | -1.69 | 136 | 0.093 | -4.556 | 2.695 | 0.021 |
| RUL R vs. xBT | R Inferior Frontal G Opercular Area, SA | 2.12 | 150 | 0.035 | 61.03 | 28.75 | 0.03 |
| | R Circular S, Superior Insula, SA | 1.84 | 150 | 0.068 | 39.526 | 21.479 | 0.022 |
| | R Inferior Frontal G Opercular Area, Mean Curv | 2.83 | 150 | 0.005 | 0.005 | 0.002 | 0.051 |
| | R Amygdala, Paralaminar Volume | -2.81 | 136 | 0.006 | -6.904 | 2.456 | 0.055 |
| | R Hippocampus, Parasubiculum Volume | -2.61 | 136 | 0.010 | -5.758 | 2.203 | 0.048 |
| | R Hippocampus Head, Molecular Layer Volume | -2.75 | 136 | 0.007 | -7.364 | 2.681 | 0.053 |
| | R Amygdala, Basal Volume | -2.54 | 136 | 0.012 | -6.129 | 2.415 | 0.045 |
| | R Hippocampus Head, CA1 Volume | -2.63 | 136 | 0.009 | -6.785 | 2.578 | 0.048 |
| | R Hippocampus Head, CA4 Volume | -2.37 | 136 | 0.019 | -6.090 | 2.567 | 0.040 |
| | R Hippocampus Head, Dentate et al. Volume | -2.21 | 136 | 0.029 | -5.514 | 2.497 | 0.035 |
| | R Amygdala, AB Volume | -1.67 | 136 | 0.098 | -3.930 | 2.357 | 0.020 |
| RUL R vs. NC | R Inferior Frontal G Opercular Area, SA | 2.17 | 150 | 0.031 | 69.23 | 31.84 | 0.03 |
| | R Circular S, Superior Insula, SA | 2.26 | 150 | 0.025 | 53.850 | 23.788 | 0.033 |
| | R Inferior Frontal G Opercular Area, Mean Curv | 2.38 | 150 | 0.018 | 0.005 | 0.002 | 0.037 |
| | R Amygdala, Paralaminar Volume | -1.69 | 136 | 0.094 | -4.563 | 2.704 | 0.021 |
| | R Hippocampus, Parasubiculum Volume | -1.28 | 136 | 0.201 | -3.116 | 2.426 | 0.012 |
| | R Hippocampus Head, Molecular Layer Volume | -1.44 | 136 | 0.152 | -4.252 | 2.952 | 0.015 |
| | R Amygdala, Basal Volume | -1.23 | 136 | 0.221 | -3.269 | 2.659 | 0.011 |
| | R Hippocampus Head, CA1 Volume | -1.23 | 136 | 0.222 | -3.482 | 2.838 | 0.011 |
| | R Hippocampus Head, CA4 Volume | -0.92 | 136 | 0.360 | -2.597 | 2.826 | 0.006 |
| | R Hippocampus Head, Dentate et al. Volume | -0.91 | 136 | 0.362 | -2.512 | 2.749 | 0.006 |
| | R Amygdala, AB Volume | -0.80 | 136 | 0.427 | -2.069 | 2.595 | 0.005 |

**Figure S1**

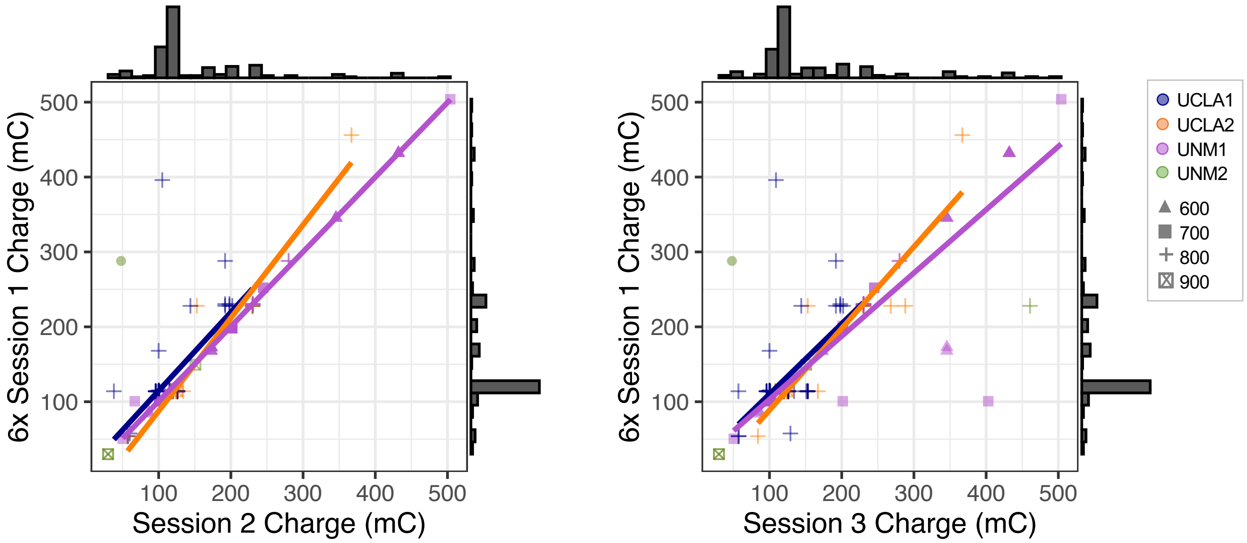


**Figure S1.** Relationships between six times (6x) ECT1 charge at seizure titration/threshold and charge during ECT2 (left) and ECT3 (right). These plots demonstrate changes in stimulation parameters across sessions for some patients. Color denotes cohort.

**Figure S2**

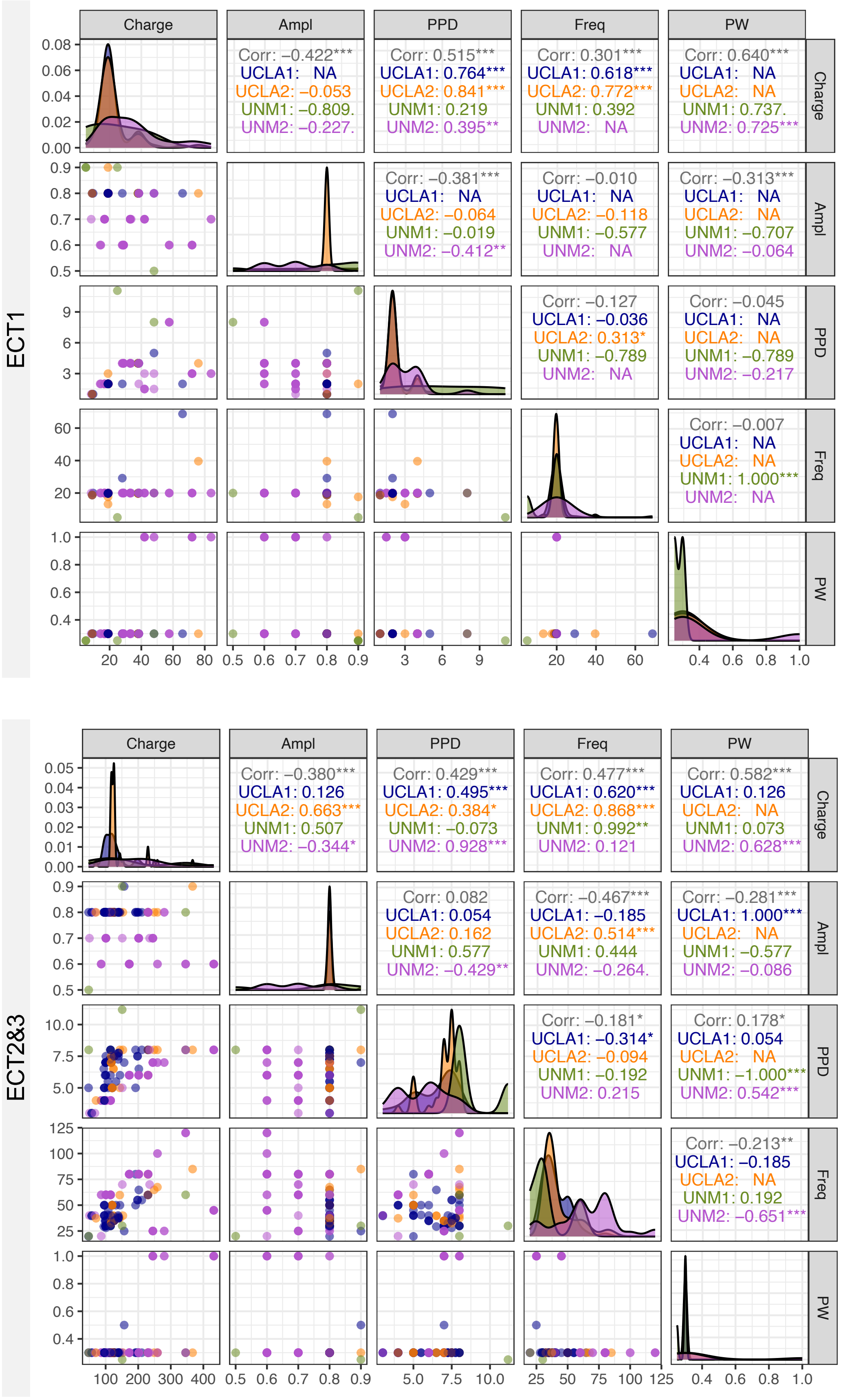


**Figure S2.** Relationships between each pair of ECT Parameters for ECT1 (top grid) and ECT2&3 (bottom grid) are are displayed. Lower left triangle of plots display scatter plots, upper right triangle display Pearson's r, and diagonal displays curved histograms for each metric. Color denotes cohort.

**Figure S3**

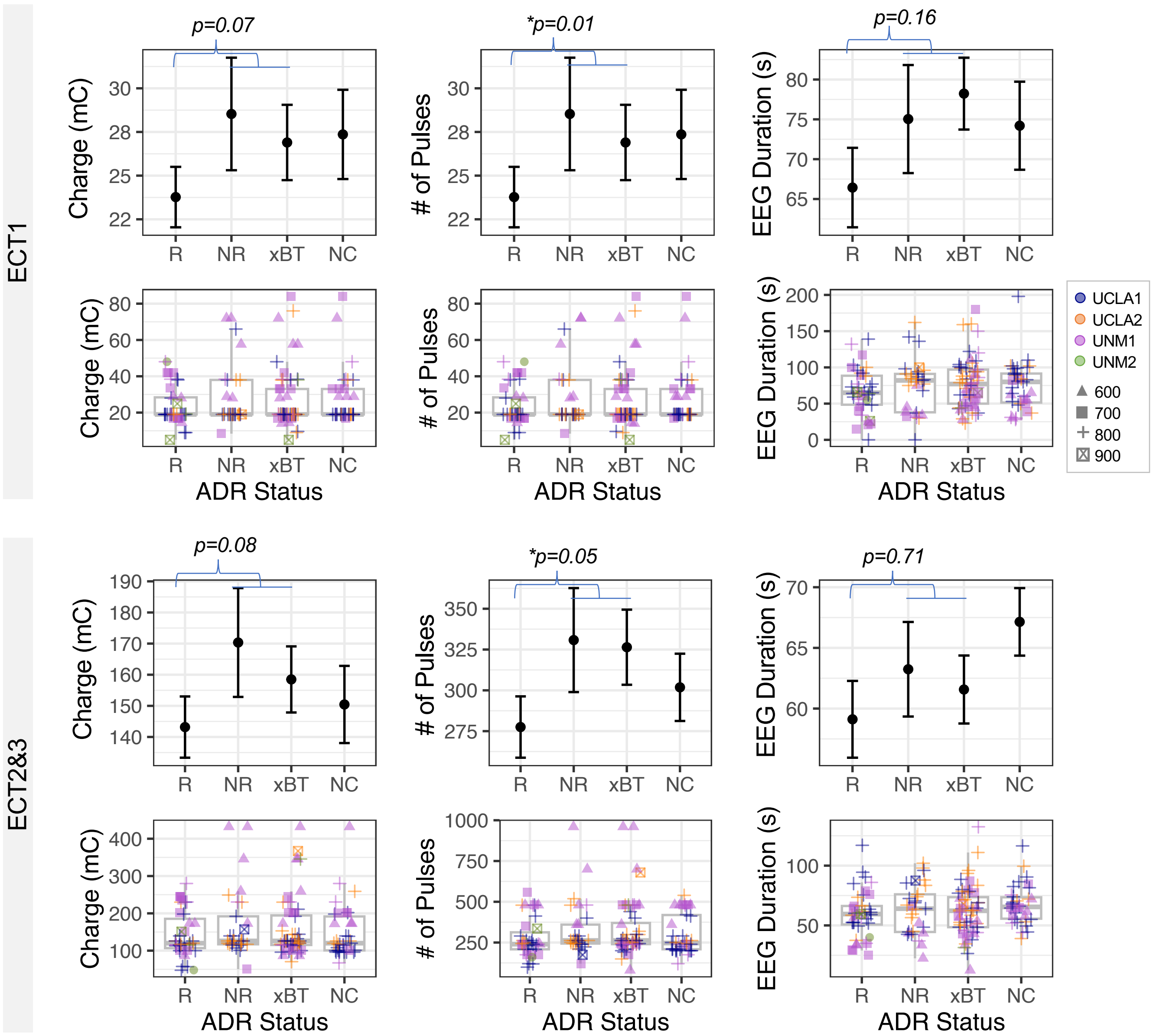


**Figure S3.** ECT Parameters for ECT1 (top 2 rows) and ECT2&3 (bottom 2 rows) are plotted against antidepressant response. In first and third rows, mean and standard error are plotted for responders (R, >50% HDRS improvement), nonresponders (NR, <50% HDRS improvement), transition to bitemporal ECT (xBT), and non-completers (NC). In second and fourth rows, boxplots are displayed, along with datapoints for each participant. Color indicates cohort, and shape reflects amplitude.
